# Genome-wide studies define new genetic mechanisms of IgA vasculitis

**DOI:** 10.1101/2024.10.10.24315041

**Authors:** Lili Liu, Li Zhu, Sara Monteiro-Martins, Aaron Griffin, Lukas J. Vlahos, Masashi Fujita, Cecilia Berrouet, Francesca Zanoni, Maddalena Marasa, Jun Y. Zhang, Xu-jie Zhou, Yasar Caliskan, Oleh Akchurin, Samhar Al-Akash, Augustina Jankauskiene, Monica Bodria, Aftab Chishti, Ciro Esposito, Vittoria Esposito, Donna Claes, Vladimir Tesar, Thomas K. Davis, Dmitry Samsonov, Dorota Kaminska, Tomasz Hryszko, Gianluigi Zaza, Joseph T. Flynn, Franca Iorember, Francesca Lugani, Dana Rizk, Bruce A. Julian, Guillermo Hidalgo, Mahmoud Kallash, Luigi Biancone, Antonio Amoroso, Luisa Bono, Laila-Yasmin Mani, Bruno Vogt, Fangming Lin, Raji Sreedharan, Patricia Weng, Daniel Ranch, Nianzhou Xiao, Alejandro Quiroga, Raed Bou Matar, Michelle N. Rheault, Scott Wenderfer, Dave Selewski, Sigrid Lundberg, Cynthia Silva, Sherene Mason, John D. Mahan, Tetyana L. Vasylyeva, Krzysztof Mucha, Bartosz Foroncewicz, Leszek Pączek, Michał Florczak, Małgorzata Olszewska, Agnieszka Gradzińska, Maria Szczepańska, Edyta Machura, Andrzej Badeński, Helena Krakowczyk, Przemysław Sikora, Norbert Kwella, Monika Miklaszewska, Dorota Drożdż, Marcin Zaniew, Krzysztof Pawlaczyk, Katarzyna Siniewicz-Luzeńczyk, Andrew S. Bomback, Gerald B. Appel, Claudia Izzi, Francesco Scolari, Anna Materna-Kiryluk, Malgorzata Mizerska-Wasiak, Laureline Berthelot, Evangeline Pillebout, Renato C. Monteiro, Jan Novak, Todd Jason Green, William E. Smoyer, M. Colleen Hastings, Robert J. Wyatt, Raoul Nelson, Javier Martin, Miguel A. González-Gay, Philip L. De Jager, Anna Köttgen, Andrea Califano, Ali G. Gharavi, Hong Zhang, Krzysztof Kiryluk

## Abstract

IgA vasculitis (IgAV) is a pediatric disease with skin and systemic manifestations. Here, we conducted genome, transcriptome, and proteome-wide association studies in 2,170 IgAV cases and 5,928 controls, generated IgAV-specific maps of gene expression and splicing from blood of 255 pediatric cases, and reconstructed myeloid-specific regulatory networks to define disease master regulators modulated by the newly identified disease driver genes. We observed significant association at the *HLA*-*DRB1* (OR=1.55, P=1.1×10^−25^) and fine-mapped specific amino-acid risk substitutions in DRβ1. We discovered two novel non-HLA loci: *FCAR* (OR=1.51, P=1.0×10^−20^) encoding a myeloid IgA receptor FcαR, and *INPP5D* (OR=1.34, P=2.2×10^−09^) encoding a known inhibitor of FcαR signaling. The *FCAR* risk locus co-localized with a cis-eQTL increasing *FCAR* expression; the risk alleles disrupted a *PRDM1* binding motif within a myeloid enhancer of *FCAR*. Another risk locus was associated with a higher genetically predicted levels of plasma IL6R. The *IL6R* risk haplotype carried a missense variant contributing to accelerated cleavage of IL6R into a soluble form. Using systems biology approaches, we prioritized IgAV master regulators co-modulated by *FCAR*, *INPP5D* and *IL6R* in myeloid cells. We additionally identified 21 shared loci in a cross-phenotype analysis of IgAV with IgA nephropathy, including novel loci *PAID4, WLS*, and *ANKRD55*.

## Introduction

IgA vasculitis (IgAV), also known as Henoch-Schönlein purpura (HSP), is the most common form of childhood vasculitis, affecting 10-27 children per 100,000 per year^1^. IgAV is characterized by IgA deposition in blood vessel walls in the skin, joints, gastrointestinal tract, and the glomerular mesangium^2,3^. IgAV can lead to serious complications, including kidney failure^4^. The exact pathogenic mechanisms of IgAV remain unclear, resulting in the general lack of targeted treatments and preventive strategies^5^. The contribution of common genetic variants to IgAV risk has not been systematically assessed, although the HLA region has been implicated in small studies^6–9^.

Herein, we report the largest genetic study of IgAV involving 8,098 individuals of European and East Asian ancestries (2,170 cases and 5,928 controls). By combining genome-wide association study (GWAS), transcriptome-wide association study (TWAS), and proteome-wide association study (PWAS) approaches, our study implicated myeloid cells as central to the disease pathogenesis, and prioritized *HLA-DRB1*, *FCAR, INPP5D,* and *IL6R* as candidate causal genes for IgAV. To assess the transcriptional consequences of the newly identified risk alleles, we performed blood RNA sequencing of 255 pediatric cases with IgAV and built a disease context-specific atlas of expression and splicing quantitative trait loci (QTLs). Using methods proposed for cancer^10,11^, we reconstructed a myeloid-specific regulatory network based on blood single cell transcriptomes and identified proteins representing candidate master regulators of IgAV-related myeloid cell state whose activity is modulated by the candidate causal genes. In aggregate, our comprehensive genetic and systems biology approach defined new pathogenic mechanisms and provided strong genetic support for novel therapeutic targets in IgAV.

## Results

### Genome-wide Association Study (GWAS)

We performed a GWAS meta-analysis of three large case-control cohorts (two newly genotyped and one previously published cohort^7^) comprising a total of 8,098 individuals (2,170 IgAV cases and 5,928 controls). The study cohorts are summarized in **Table S1** with detailed descriptions provided in the **Methods.** In each cohort, the controls were recruited from the same ancestral populations and geographic regions as the cases and had no history of kidney or autoimmune diseases. Of the three cohorts, two were of European ancestry (1,025 cases and 3,173 controls), and one cohort was of East-Asian ancestry (1,145 cases and 2,755 controls). All cohorts were genotyped with high-density SNP arrays, ancestrally matched using principal component-based methods^12^, and imputed using ancestry-specific genome sequence reference panels^13^ (**Fig. S1A-D**). In the meta-analysis, we observed negligible genomic inflation, consistent with the absence of systematic biases (λ=1.02, **Fig. S1E**). We detected three genome-wide significant loci (P<5×10^−8^, **Fig. 1**), including the *HLA* locus (OR=1.55, P=1.1×10^−25^) and two novel non-HLA loci, on chr.2q37.1 (*INPP5D*, OR=1.34, P=2.2×10^−09^) and chr.19q13.42 (*FCAR*, OR=1.51, P=1.0×10^−20^). Conditional analysis identified two independent signals within the HLA region, but no secondary signals at the non-HLA loci (**Table S2**). We also detected 14 suggestive loci based on P<1×10^-5^, including the *TNFSF13* and *IL6R* loci (**Table S3**).

**Figure 1.**
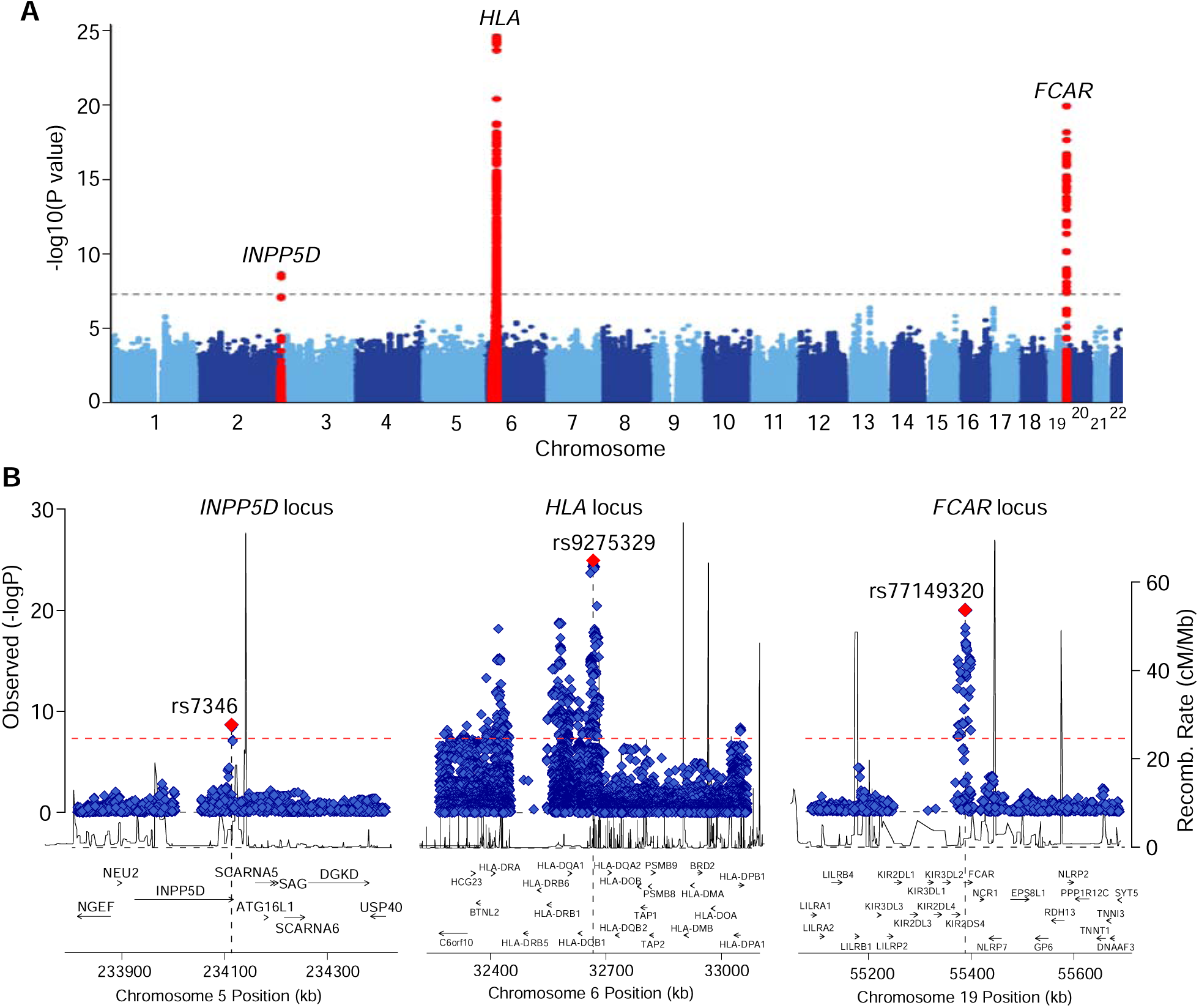
GWAS meta-analysis for IgAV identified 3 genome-wide significant regions. **(A)** Manhattan plot with genome-wide significant loci highlighted in red; the dotted horizontal line indicates a genome-wide significance threshold (P=5×10-8). **(B)** Regional plots for the *INPP5D* locus (left), the *HLA* locus (middle), and the *FCAR* locus (right). The x-axis shows the physical position in kilobases (kb, hg19 coordinates) and includes known gene annotations; the left y-axis presents -log10 p-values for association statistics and the right y-axis shows the recombination rates; the dotted horizontal line indicates a genome-wide significant threshold of 5×10^−8^.

### Transcriptome-wide Association Study (TWAS) and Proteome-wide Association Study (PWAS)

TWAS^14^ and PWAS^15^ represent powerful approaches to identify causal genes for complex traits by testing the effects of genetically determined gene or protein expression on a disease risk. Our PWAS (see **Methods**) identified a single proteome-wide significant signal for soluble IL6R, with higher genetically determined plasma levels associated with greater disease risk (P=1.6×10^−6^, **Fig. 2A, Supplementary Data 1**). TWAS provided no signals of transcriptome-wide significance although the *IL6R* signal was suggestive in blood and small intestine tissues (**Supplementary Data 2**).

**Figure 2.**
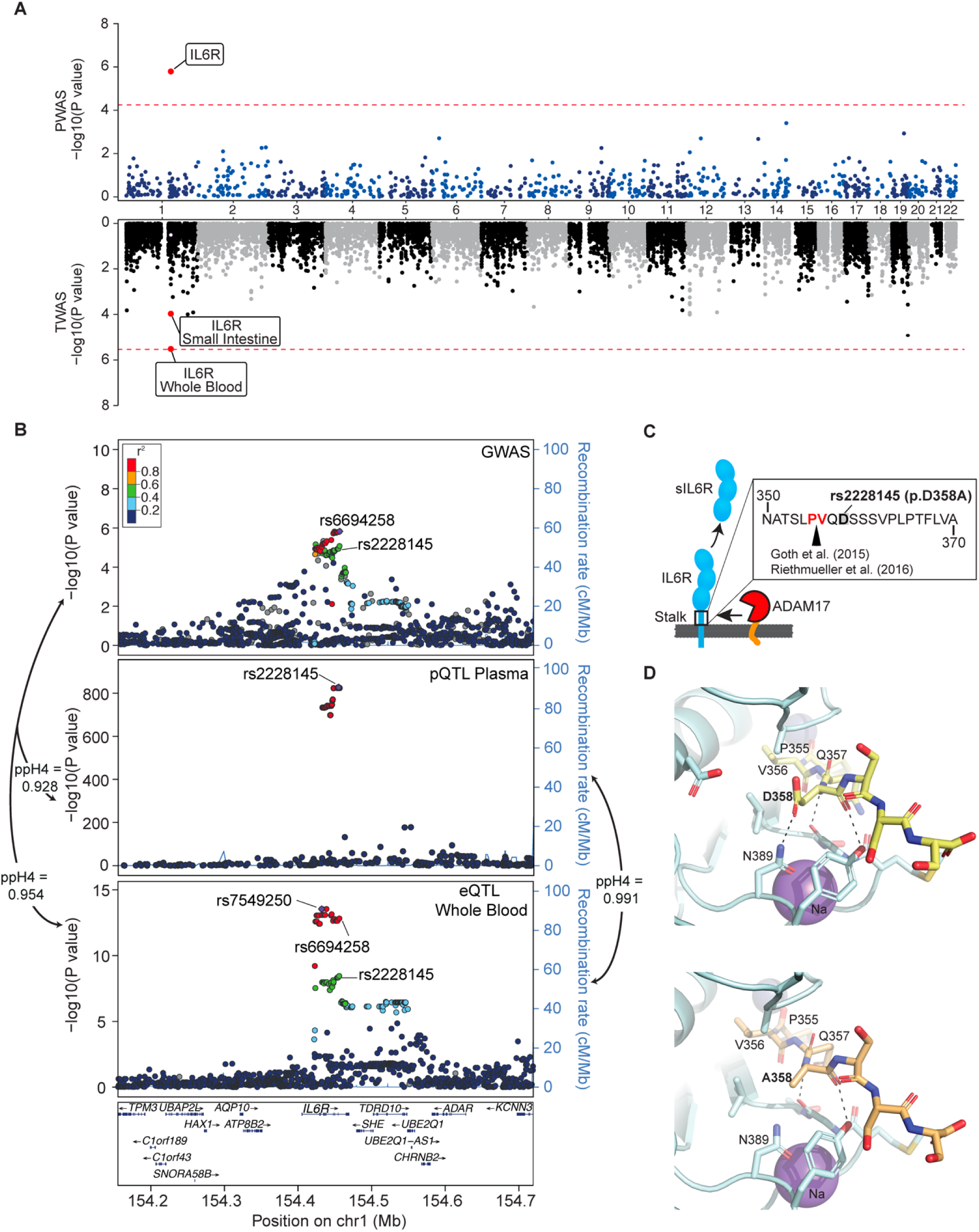
Effects of the *IL6R* locus. **(A)** Miami plot for PWAS and TWAS analyses. The red lines indicate the Bonferroni adjusted significance thresholds (5.6×10^−5^ for PWAS and 2.5×10^−6^ for TWAS). **(B)** Regional association plots for the *IL6R* locus. SNPs are plotted by position (hg19, 250kb window) versus -log10(P-values) from GWAS of IgAV (top), pQTL of plasma sIL6R levels (middle) and eQTL of *IL6R* expression in whole blood (bottom). The purple diamond highlights the most significant SNP for each association. SNPs are color-coded to reflect their LD with this SNP using LocusZoom with HapMap CEU reference. The missense variant *IL6R* p.Asp358Ala (rs2228145) is indicated in each plot. **(C)** The p.Asp358Ala substitution in relation to the cleavage site of the membrane bound IL6R. **(D)** Structure of the ADAM17 catalytic domain/IL-6R complex. The structure of ADAM17 (pale cyan ribbon render) was modeled with bound peptide 355-PVQDSSS-361 (yellow sticks in upper panel) or 355-PVQASSS-361 (pale orange sticks in lower panel) corresponding to a segment of the stalk region of IL-6R including the cleavage site (P355/V356). Residues of ADAM17 within 5 angstroms of D358 or A358 are shown as sticks. Polar interactions between residue 358 and ADAM17 are noted by dashed lines. The change D358A eliminates the interaction with N389 of ADAM17, potentially reducing the affinity of IL-6R for ADAM 17. Zinc and Sodium ions are shown as slate and purple spheres, respectively. Images generated with PyMOL (Molecular Graphics System, Version 2.0, Schrödinger, LCC).

### HLA-DRB1 Locus

Using ancestry-specific reference panels, we imputed classical HLA alleles and individual amino-acid polymorphisms at the HLA class I and II genes (**Methods**). In the East Asian cohort, gene-based analysis of classical HLA alleles demonstrated the strongest association at *HLA-DRB1* (P=1.5×10^−16^, **Fig. 3A, Table S4**). Conditioning the association on *DRB1* classical alleles eliminated the entire HLA signal (**Fig. 3A**). Among the polymorphic sites in DRβ1, position 11 was most significant (P=2.9×10^−15^) and accounted for the entire signal in the conditional analysis (**Fig. 3C** and **Table S5**). Specifically, valine at DRβ1 position 11 conveyed the strongest risk (OR=1.65, P=3.4×10^−12^, **Table S6**). This substitution tags the *HLA-DRB1*04* classical allele (also genome-wide significant, OR=1.62, P=3.9×10^−10^). In contrast, proline at DRβ1 position 11, tagging *HLA-DRB1*1501*, conveyed a protective effect (OR=0.73, P=4.5×10^−6^, **Table S6**). The same residues with concordant effects have been previously reported in the GWAS for IgA nephropathy (IgAN)^16^, suggesting shared HLA susceptibility between IgAV and IgAN.

**Figure 3.**
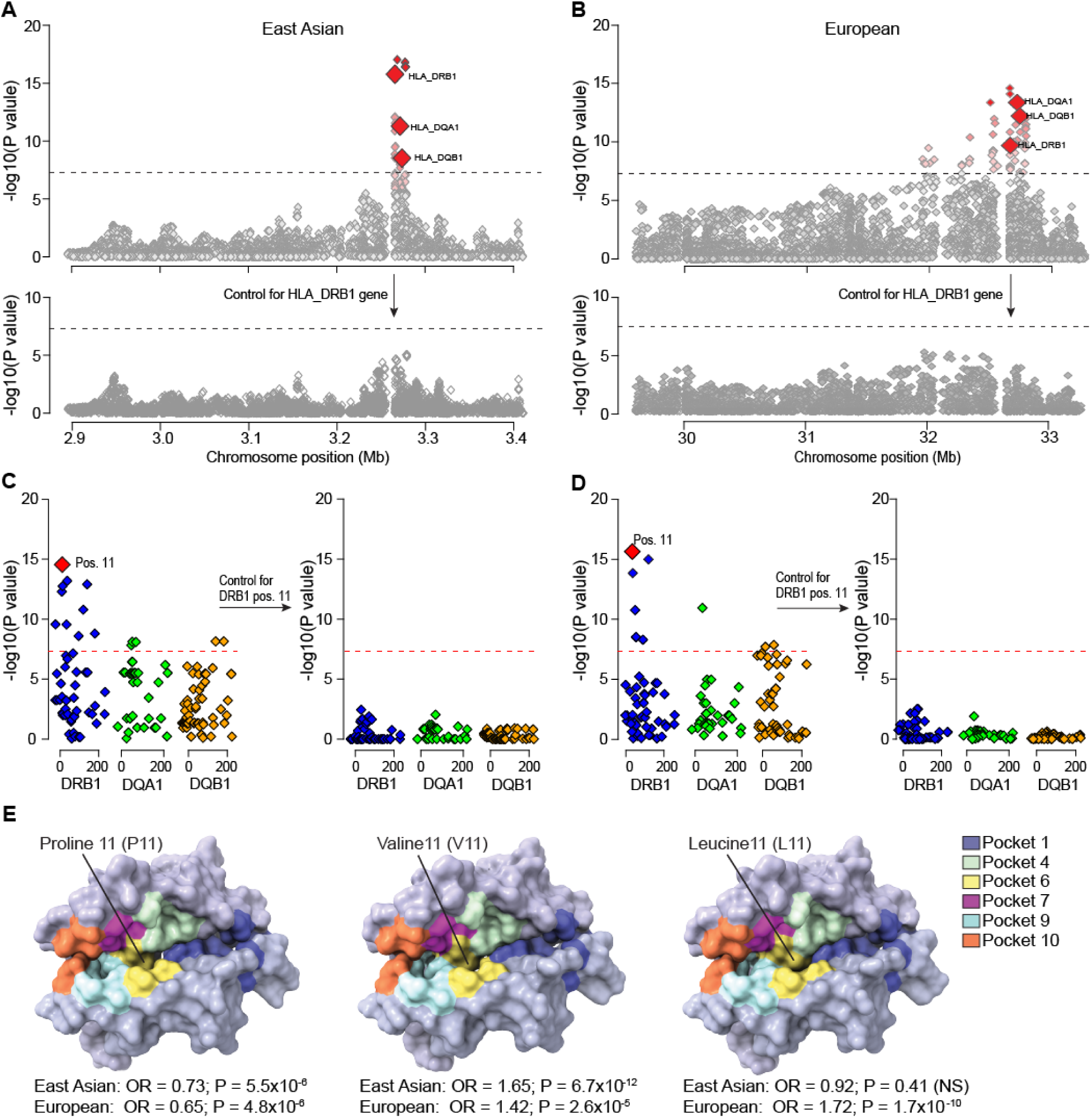
Fine mapping of the *HLA* region in East Asian and European ancestry cohorts. **(A)** Regional plot for the East Asian cohort with bi-allelic and multi-allelic association statistics for all imputed variants and classical HLA alleles; the strongest association signal was for *HLA-DRB1* (upper panel); the results for *HLA-DQA1* and *HLA-DQB1* are highlighted for reference; after controlling for *HLA-DRB1* classical alleles (black arrow), there was no residual association across the entire 3-Mb region (lower panel). **(B)** Regional plot for the European cohorts with bi-allelic and multi-allelic association statistics for all imputed variants and classical HLA alleles; after controlling for *HLA-DRB1* classical alleles, there were no significant associations in the region (lower panel). The dotted horizontal line indicates the genome-wide significance threshold (α=5×10^−8^). **(C)** East Asian analysis of polymorphic amino acid positions within DRβ1 (blue), DQα1 (green) and DQβ1 (orange) using conditional-haplotype tests; the horizontal dash line indicates the genome-wide significance level. The most strongly associated polymorphic site was position 11 in DRβ1 (left panel); after controlling for this position, there was no residual association (right panel). **(D)** European analysis of polymorphic amino acid positions within DRβ1 (blue), DQα1 (green) and DQβ1 (orange); consistent with East Asian analysis, the DRβ1 amino acid position 11 (left panel) provided the strongest signal; no additional independent positions were found upon conditioning on this position (right panel). **(E)** Structure of the DRα1-DRβ1 complex. The protective variant (P11, left) and risk variants (V11, middle and L11, right) are shown in surface model viewed into the peptide binding groove. Models are colored in a background of light blue shade, with individual peptide amino acid binding pockets shaded according to the provided color key.

Consistent with the East Asian cohort analysis, *HLA-DRB1* was also genome-wide significant in European cohorts (**Fig. 3B, Table S7**). Similarly, the analysis of polymorphic sites identified the strongest association of DRβ1 position 11, with no other independent signals after conditioning on position 11 (**Fig. 3D** and **Table S8**). We observed that leucine at position 11 was a strong risk substitution in Europeans (OR=1.72, P=1.7×10^−10^**, Table S6**). This substitution is rare in East Asian (control freq. 0.03) compared to European populations (control freq. 0.11), explaining ancestry-specificity of this signal. Consistent with the findings in East Asians, we also replicated the risk effect of valine (OR=1.42, P=2.6×10^−5^) and the protective effect of proline (OR=0.65, P=4.8×10^−6^) in Europeans.

Next, we performed detailed analysis of the potential impact of the associated amino acid risk residues on the structure and potential function of DRβ1. Notably, valine at position 11 is in tight LD with histidine at position 13 (r^2^=0.99) and tyrosine at position 96 (r^2^=0.98) forming a distinct DRβ1 risk haplotype (V11-H13-Y96). The second (European-specific) DRβ1 risk haplotype (L11-F13-E96) is formed by leucine at position 11, in LD with phenylalanine at position 13 (r^2^=0.99) and glutamic acid at position 96 (r^2^=0.99). The positions 11 and 13 map to the P6 and P4 pockets of the DRβ1 antigen-binding groove, while the position 96 is at the surface-exposed on the DR-β2 domain (**Fig. S2 and S3**). Comparisons of protein structures between the protective and risk haplotypes showed two features. First, the crevasse between pockets 6 and 4 with the protective haplotype (**Fig. S2B left**) is less accessible compared to the two risk haplotypes (**Fig. S2B middle and right**). Second, the protective substitution with arginine at position 13 provides a distinct electronegative character to the binding groove (**Fig. S2C left**). These landscape and electrostatic changes may alter which peptides are accommodated by DRβ1^17^. Moreover, the position 96 is involved in the DRβ1 interactions with the HLA-DM complex in the thymic antigen presenting cells^18^, and thus substitutions at this position may additionally alter the presentation of self-antigens for clonal deletion^19,20^ (**Fig. S3**). As shown in **Fig. S3B1-3**, residue 96 is in the binding-interface with HLA-DM β-chain. All three residues Q96 (protective), E96 and Y96 (both risk) showed differing degrees of association with DM and the aqueous environment. As this is one of several residues that contribute to the interface with DM, future studies will be needed to define the mechanism of this association. Additionally, HLA-DO, a nonclassical MHC-II protein that appears to promote tolerance to self-antigens^21,22^, binds to HLA-DM through this same interface to competitively inhibit interaction with DRβ1^23^. Thus, residues such as 96 could modulate interaction of HLA-DM with other factors involved in antigen presentation.

### INPP5D Locus

The lead SNP (rs7346) at chr.2q37.1 locus is a synonymous variant in *INPP5D* (encoding Inositol Polyphosphate-5-Phosphatase D, a.k.a. SHIP-1) and is in tight LD with rs9247 (r^2^=0.99), a missense variant p.His1157Tyr in *INPP5D* **(Table S9)**. Although there is no clear deleterious prediction for this substitution by PolyPhen2, CADD and REVEL, this variant is located within a compositionally biased region of SHIP-1 that is enriched in charged residues, and the substitution of histidine (positively charged) with tyrosine (neutral) may alter its binding energy. SHIP-1 is expressed in myeloid cells, where its activation is mediated by tyrosine phosphorylation. Once phosphorylated, SHIP-1 functions as a negative regulator of FcαR signaling, inhibiting myeloid cell proliferation, chemotaxis, and degranulation^24,25^. Phosphorylation of SHIP-1 also occurs by simultaneous engagement of IgA and IgE Fc receptors, suppressing allergic responses^26^. In mice, deletion of *Inpp5d* leads to autoinflammatory disease with neutrophilia and gastrointestinal involvement^27,28^. In GWAS, this locus has previously been associated with allergy^29,30^, and increased neutrophil^31^, eosinophil^32,33^, and monocyte counts^31^. In our meta-PheWAS across major biobanks involving 786,138 individuals (**Fig. S4A and Supplementary Data 3**), the IgAV-risk allele at this locus was additionally associated with hypothyroidism (OR=1.05, P=2.5×10^−10^) and dermatitis herpetiformis (a skin rash due to IgA response against epidermal transglutaminase and involving FcαR-mediated neutrophil degranulation^34^, OR=1.95, P=8.7×10^−7^).

### *FCAR* Locus

The index SNP at 19q13.42 (rs77149320) maps to the intronic region of *FCAR*, encoding IgA receptor FcαR (CD89), a transmembrane glycoprotein expressed specifically on myeloid cells. FcαR interacts with IgA immune complexes triggering phagocytosis, cytotoxicity, and release of inflammatory mediators. In prior GWAS, this locus has been associated with IgAN^16^ and inflammatory bowel disease^35^. In our meta-PheWAS, the most significant association of this locus was with higher risk of “nephritis and nephropathy with pathological lesion” (**Fig. S4B)**. The index SNP is in tight LD (r^2^>0.8) with seven variants intersecting a putative cis-regulatory region in myeloid cells defined by ABC models^36^, confirmed by FUN-LDA functional scores^37^, and intersecting open chromatin by monocyte-specific ATAC-seq in 25 healthy donors^38^ (**Fig. 4A, Tables S10**). Two of the seven top risk alleles in LD (rs4806602-G and rs73065472-T) disrupt the predicted binding motif for PRDM1^39^, a transcriptional repressor triggered by infections via Toll-like receptor and STAT signaling (**Fig. 4A**). PRDM1 activates NF-κB/TNF-R signaling by repressing NLRP2^40^, maintains effector functions of BD and TDcells^40,41^, and promotes myeloid cell differentiation^41^. In agreement with these findings, our network-based differential activity analysis demonstrated higher regulatory activity of PRDM1 in IgAV cases versus controls (FDR=0.0037, see below).

**Figure 4.**
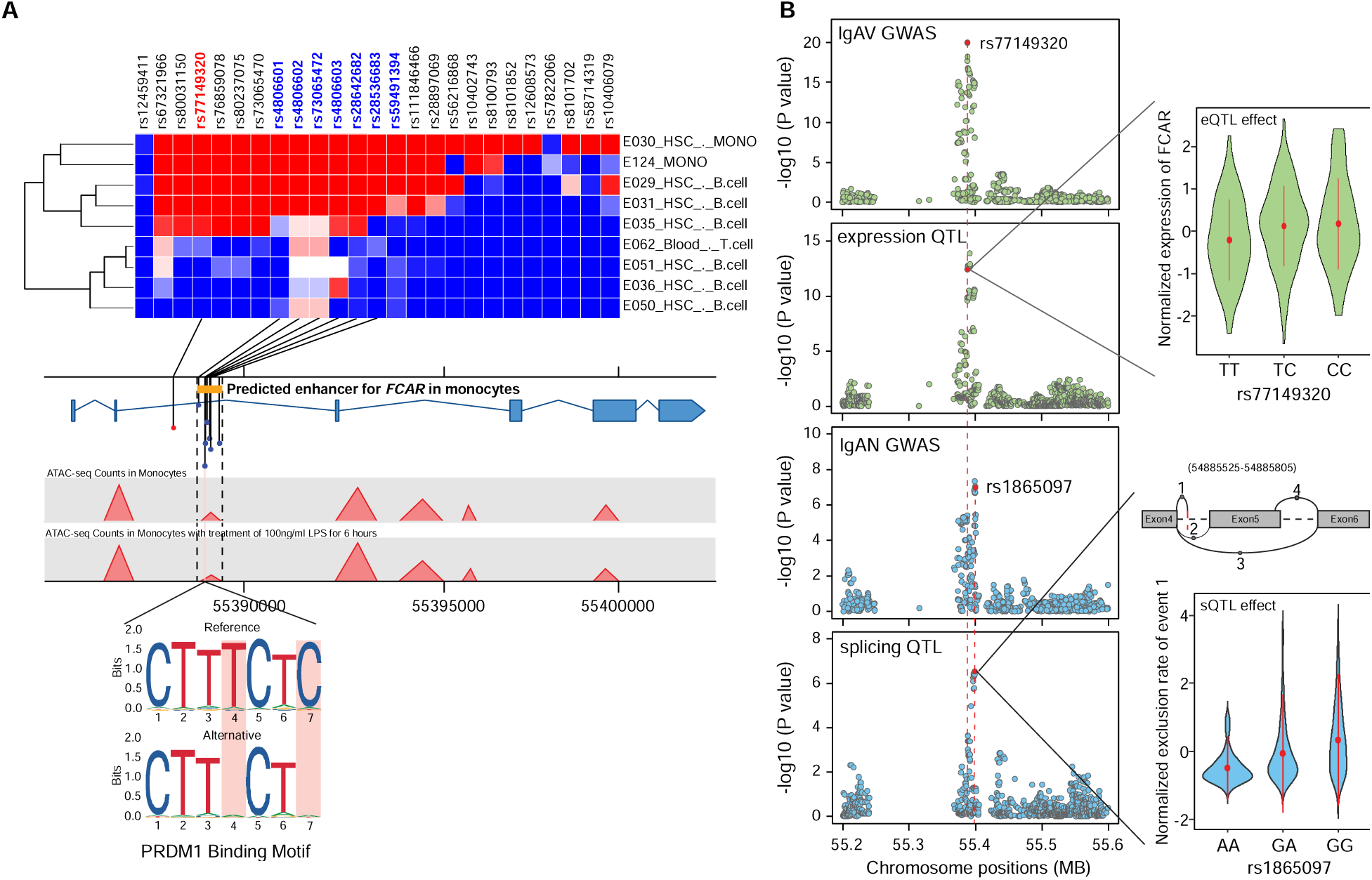
Functional annotations of the *FCAR* locus. (a) The heatmap of FUN-LDA functional scores for the lead SNP (rs77149320, red) and its high LD proxies (r^2^>0.8, only SNPs and cell types with positive scores are depicted). The 7 SNPs in LD (names in blue) intersected an ABC model-predicted intronic enhancer of *FCAR*. This regulatory region was further confirmed by an ATAC-seq peak in monocytes before and after LPS treatment. The prioritized SNPs were analyzed against transcription factor binding motifs predicted by JASPAR, and two risk alleles (rs4806602-G and rs73065472-T in perfect LD and two base pairs apart) were found to disrupt the PRDM1 binding motif; (b) Co-localization of the *FCAR* locus between GWAS for IgAV and blood eQTL for *FCAR* in IgAV (top two panels), and GWAS for IgAN and blood sQTL for *FCAR* in IgAV (bottom two panels). The eQTL effect shown in the right upper panel (x-axis: genotypes, y-axis: normalized expression of *FCAR*). The sQTL effect shown in the right bottom panel (x-axis: genotypes, y-axis: normalized exclusion rate of chr19:54885525-54885805 region based on hg38 annotations, the splice model depicted above).

### *IL6R* Locus

Soluble IL6R (Interleukin 6 receptor) reached a proteome-wide significance in PWAS and was supported by a suggestive signal in GWAS. Interestingly, the top GWAS risk allele (rs6694258-A, OR=1.21, P=1.49×10^−6^, **Table S3**) was associated with higher plasma protein levels of IL6R, but lower *IL6R* transcript levels of in blood (**Fig. 2B**). The GWAS locus co-localized with both expression QTL (posterior probability 95%) and protein QTL (posterior probability 93%) supporting a shared casual haplotype (**Fig. 2B and Supplementary Data 4**). Four SNPs in strong LD with rs6694258 (r^2^≥0.8) intersected *IL6R* enhancers predicted by ABC model^36^ and confirmed by ATAC-seq in monocytes^38^ (**Fig. S5** and **Table S10**). The observed protein QTL effect of this locus was particularly strong, with the top SNP (rs4129267) alone explaining 25.9% of variance in plasma IL6R levels. The second top variant (rs2228145, r^2^=0.998 with rs4129267), is a missense variant resulting in p.Asp358Ala substitution in the stalk region, close to the ADAM17 cleavage site of IL6R (between position 355 and 356; **Fig. 2C**)^42^, and demonstrated to increase sIL6R levels in prior studies^43–45^. We modeled IL6R-ADAM17 structures based on published studies^46^ to demonstrate that Asp358 forms a hydrogen bond with Asn389 of ADAM17, whereas p.Asp358Ala substitution in IL6R results in the loss of this interaction (**Fig. 2D** and **Fig. S6**). This may decrease affinity between IL6R and ADAM17, potentially allowing for faster release upon cleavage and higher turnover rate. Prior studies showed that a peptide harboring the Asp358Ala substitution is cleaved more rapidly by ADAM17 than the peptide with the Asp358 residue^45^. In our Meta-PheWAS of rs2228145, the risk allele for IgAV was additionally associated with a higher risk of asthma and urticaria, and a reduced risk of atrial fibrillation/flutter and coronary syndrome (**Fig. S4C and Supplementary Data 5**).

### Shared and distinct loci with IgAN

To examine the genetic relationship between IgAV and IgAN, we tested genome-wide genetic correlation using results from the latest GWAS for IgAN^16^. We observed a significant positive genetic correlation between IgAV and IgAN (r_g_=0.77, P=2.8×10^−12^) also present after excluding the HLA region (r_g_=0.69, P=0.016). To better understand the shared and distinct loci between IgAN and IgAV, we performed a cross-phenotype GWAS (see **Methods**). We observed 21 genome-wide significant loci including 3 novel loci, near *PAID4, WLS*, and *ANKRD55* (**Fig. S7, Table S11**). All three IgAV-risk loci (*HLA, INPP5D*, and *FCAR* loci) became more significant in the cross-phenotype analysis (**Fig. S7**), but at the *FCAR* locus significant heterogeneity was detected between IgAV and IgAN (I^2^=94.98, p<0.0001). While the *INPP5D* locus perfectly co-localized between the two traits (posterior probability of shared causal variant 0.99), the *FCAR* locus did not colocalize (posterior probability of different causal variants 0.98, **Table S12)**, implying distinct genetic mechanisms between IgAV and IgAN at this locus.

### Maps of blood expression and splicing QTLs in IgAV

To better interpret the effects of IgAV-risk alleles, we performed blood RNA-seq in a cohort of 255 IgAV cases and generated genome-wide maps of disease-context specific expression and splicing QTLs. In total, we identified 7,594 cis-eGenes (11,597 independent eQTLs) at FDR<0.05 (**Supplementary Data 6 and 7**). Among them, 1,345 eGenes were IgAV context-specific when compared to GTEx v7 whole blood dataset (**Fig. S8**). These IgAV-specific eGenes were enriched in immune-related pathways and mouse phenotypes, including TNF and interleukin signaling (**Fig. S8**). We also identified 1,192 sGenes (**Supplementary Data 8**), including 226 IgAV-specific sGenes enriched in the biological processes related to immunity, such as leukocyte and lymphocyte activation and regulation of immune system responses (**Table S13**). The eQTL and sQTL results were then used to investigate the consequences of IgAV-risk alleles.

Based on these maps, we observed strong colocalization between the *FCAR* locus for IgAV and *FCAR* cis-eQTL (posterior probability of shared causal variant of 0.96). IgAV-risk allele was associated with increased blood *FCAR* mRNA expression (**Fig. 4B**). Differential gene expression analysis confirmed higher *FCAR* gene expression in IgAV cases compared to controls (fold change=1.3, P=6.84×10^−06^). In contrast, the distinct *FCAR* association signal for IgAN did not co-localize with neither eQTL nor GWAS for IgAV, but it did co-localize strongly with an sQTL for *FCAR* (posterior probability of shared causal variant of 0.99). One of the top IgAN-risk alleles (rs1865097-A) is a deep intronic variant associated with lower exclusion rate of the chr19:54885525-54885805 region (**Fig. 4B**). This variant, located at the end of the alternative splicing event (chr19:54885805) functions as a cryptic splice acceptor site. The protective exclusion of chr19:54885525-54885805 region results in the transcript excluding exon 5 of *FCAR* encoding the transmembrane domain.

### IgAV transcriptional regulators in myeloid cells

Given that all identified candidate causal genes are expressed specifically in a myeloid cell lineage, we generated a myeloid-specific transcriptome-wide regulatory gene network based on blood single-cell RNA-seq data from 163 reference individuals (**Figs. 5, S9 and S10, and Methods**). To overcome the sparseness of single-cell gene expression matrix, we created a pseudo-bulk gene expression profile for the myeloid cell cluster. We then used a mutual-information-based network reconstruction method, ARACNe3, to identify 812,995 significant pairwise regulatory relationships between 2,212 transcriptional regulators and their targets. Next, we generated the differential gene expression signature using the bulk RNA-seq data from our dataset of 255 IgAV patients and 38 healthy controls controlling for age, sex, genetic ancestry, and CIBERSORTx^47^-deconvolved cell fractions (**Fig. 5B**). Based on the myeloid-specific ARACNe3 regulons (**Fig. 5C**) and the adjusted differential gene-expression signatures, we inferred differentially activated regulators in IgAV cases compared to controls using the NaRnEA algorithm^48^. This analysis identified 773 regulators with increased activity and 916 with decreased activity in IgAV cases compared to controls based on FDR<0.05 (**Fig. 5D, and Supplementary Data 9**). The regulators activated in the disease state were significantly enriched in the TGFβ signaling pathway (**Table S14**). We have previously identified genetic support for the involvement of transcriptional regulators *LITAF*, *HDAC7*, *RUNX2*, and *RUNX3* in the determination of IgA levels^49^. In IgAV cases, both *LITAF* and *HDAC7* were inferred to have increased activity in myeloid cells (FDR=1.5×10^−10^ and 4.1×10^−8^, respectively), while *RUNX2* and *RUNX3* to have suppressed activity compared to controls (FDR=1.5×10^−3^ and 5.9 ×10^−14^, respectively). These observations are direction-consistent with the previously reported genetic effects of these regulators on IgA levels^49^, supporting global activation of the regulatory programs that increase IgA production in IgAV.

**Figure 5.**
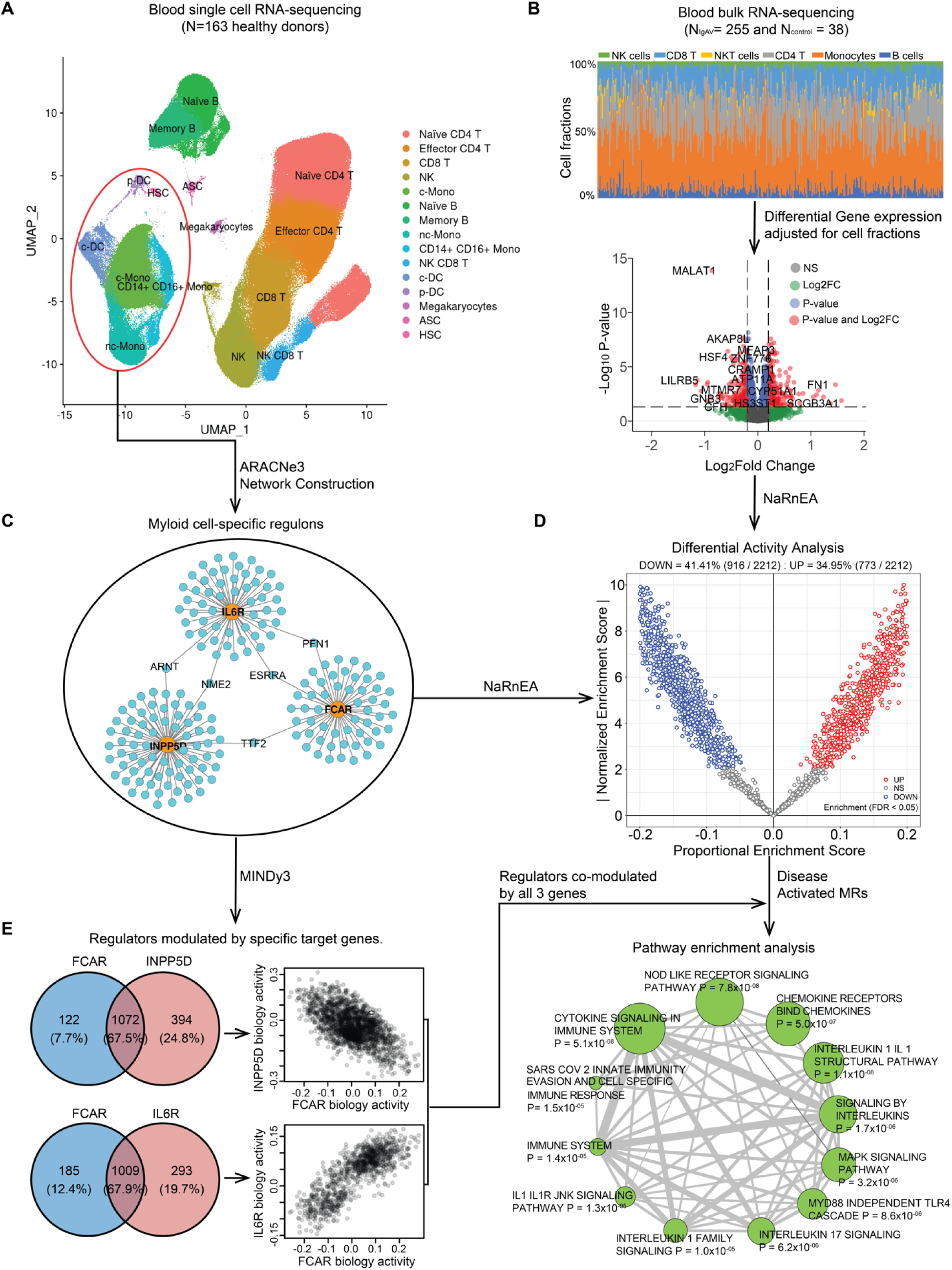
Master regulators of IgA vasculitis modulated by the GWAS candidate genes. (a) the UMAP plot for the reference single cell RNA-seq data from 163 healthy individuals. The blood cells were clustered and annotated into 15 different immune cell types highlighted in different colors; (b) Deconvolution and differential gene expression analysis based on blood bulk RNA-seq data from 255 IgAV cases and 38 health controls. The upper panel shows the distribution of different cell type fractions across all 293 individuals. The volcano plot (lower panel: x-axis denotes the log2 Fold Change and the y-axis indicates the adjusted P values) with differentially expressed genes in red (P_adjusted_<0.05 and log2 Fold Change>0.2) after adjusting for cell type fractions; (c) The myeloid regulons reconstructed transcriptome-wide based on the myeloid lineage cells (circled in red above) using ARACNe3 software (a subset of only 3 regulons shown to illustrate this step). (d) Differential activity analysis based on the myeloid regulons and cell fraction-adjusted differential gene expression signature in IgAV cases compared to controls. The x-axis indicates the proportional enrichment score (effect size for gene set enrichment that was normalized by set’s size, parameterization, and gene expression signature). The y-axis shows the normalized score compared to the expectation under the null hypothesis. The red and blue colors indicate the regulators with higher and lower activity, respectively, in IgAV cases compared to healthy controls. (e) Regulators co-modulated by the GWAS candidate genes: 1,072 regulators co-modulated by *FCAR* and *INPP5D* with mostly opposed effects on their biological activities (upper panel) and 1,009 regulators co-modulated by *FCAR* and *IL6R* with direction consistent effects on their biological activities (lower panel). The targets of the top master regulators co-modulated by all 3 genes were enriched in multiple immune related pathways based on FDR< 0.05 (depicted as enrichment map with a node size proportional to the statistical significance of enrichment and edge thickness proportional to gene set overlaps).

### Master regulators modulated by IgAV candidate genes

Next, we hypothesized that *FCAR, INPP5D,* and *IL6R* transcripts may modulate the activity of proteins representing Master Regulators (MRs) of IgAV-related myeloid cell state. To test this hypothesis, we used MINDy^50^, a new algorithm that identifies potential modulators of transcriptional interactions by analyzing statistical dependencies between a transcription factor and its targets conditioned on the expression level of a candidate modulator. By applying this approach to all myeloid regulators using GWAS candidate genes as potential modulators and myeloid-specific ARACNe3 regulons, we identified 1,194 regulators modulated by *FCAR* (**Supplementary Data 10**), 1,466 by *INPP5D* (**Supplementary Data 11**), and 1,302 by *IL6R* (**Supplementary Data 12**) at FWER<0.05 (**Fig. 5E**). A total of 1,072 regulators were co-modulated by both *FCAR* and *INPP5D* with a strong negative correlation in biological activity, indicating that the regulator activities enhanced by *FCAR* were generally repressed by *INPP5D* (Spearman’s r=-0.52, P<2.2×10^−16^). This phenomenon is fully consistent with the established role of *INPP5D* as a negative regulator of FcαR signaling. Moreover, a total of 1,009 regulators were co-modulated by *FCAR* and *IL6R*, with strong positive correlation of their biological activity (Spearman’s r=0.78, P<2.2×10^−16^), suggesting concordant effects on gene regulation in myeloid cells. Lastly, to identify MRs affected by our genetic risk loci, we intersected the 50 most activated and 50 most inactivated IgAV MRs with the genes co-modulated by *FCAR, INPP5D,* and *IL6R* transcripts. This nominated 28 high-priority MRs (**Table S15**), targets of which were significantly enriched in key immune-related pathways, including cytokine, chemokine, and Nucleotide-binding oligomerization domain (NOD)-like receptor signaling pathways (**Fig. 5E**).

## Discussion

By combining hypothesis-free genome, transcriptome, and proteome-wide association approaches with gene expression and splice QTL analyses and network-based analyses, our study uncovered new genetic mechanisms of IgAV. We fine-mapped *HLA-DRB1*, *FCAR*, *INPP5D*, and *IL6R* as the candidate causal genes for IgAV and characterized their effects on the disease-related regulons in myeloid cells. We also demonstrated shared genetic susceptibility between IgAV and IgAN and extensively characterized the shared and distinct susceptibility loci in combined and separate analyses of both conditions.

Our fine-mapping studies revealed that the HLA signal was largely explained by the polymorphic position 11 in DRβ1, which is in tight LD with additional substitutions at positions 13 and 96. Our haplotype-based analyses defined a protective haplotype (P11-R13-Q96) and two risk haplotypes (L11-F13-E96 and V11-H13-Y96). While positions 11 and 13 mapped to the surface of pockets 4 and 6 of peptide binding groove of DRβ1 potentially changing antigen binding affinity, position 96 may be involved in the interaction between DRβ1 and HLA-DM/DO, potentially affecting antigenic peptide loading to DRβ1. Notably, the same haplotypes have a direction-consistent effect in IgAN, suggesting that the same antigen presentation defect is shared by both conditions.

Two novel non-HLA risk loci, *FCAR* and *INPP5D*, converge on the same pathway in myeloid cells, highlighting the central role of the FcαR (CD89) signaling in the pathogenesis of IgAV. In prior studies, elevated serum levels of IgA1-CD89 complexes have been reported in children with IgAV^51^ and IgAN^52^. However, our results demonstrate distinct genetic mechanisms underlying genetic associations of the *FCAR* locus in IgAV versus IgAN. In IgAV, the risk alleles disrupt a binding motif of *PRDM1* in a monocyte-specific *FCAR* enhancer and are associated with increased *FCAR* gene expression. In IgAN, the risk alleles are associated with a lower rate of a splicing event that excludes a transmembrane domain of FcαR, leading to increased expression of the full-length isoform.

In contrast, the risk variant in *INPP5D* encoding SHIP-1, a known negative regulator of *FCAR* signaling^25,26^, had the same effect in IgAV and IgAN. The inhibition of SHIP-1 is known to generate molecular profiles consistent with activation of inflammasome^53^ and increased phagocytosis^54,55^. We hypothesize that p.His1169Tyr substitution may disrupt the function of SHIP-1, but further studies are needed to understand the precise effect of this variant.

Our study demonstrated that the PWAS approach could effectively implicate a new causal gene when GWAS by itself is underpowered to establish a significant association. The PWAS signal for soluble IL6R in IgAV was supported by a colocalization between GWAS and protein QTL. About 85% of circulating sIL6R is generated by proteolysis^56^, and one of the top protein QTL variants was p.Asp358Ala, a substitution mapping to the proteolytic cleavage site in the stalk region of IL6R^57^, and previously associated with elevated serum levels of sIL6R^58^ and reduced membrane-bound IL6R^59,60^. Once cleaved, sIL6R can interact with circulating IL6 and activate gp130, a signal transducer expressed on multiple cell types, resulting in systemic pro-inflammatory effects^58–60^. In aggregate, these results support a pathogenic role of sIL6R and suggest an opportunity to repurpose Tocilizumab, a monoclonal antibody blocking IL6R, as a potential new treatment for IgAV.

We additionally characterized the pattern of global transcriptional dysregulation in circulating immune cells of children with IgAV. Our methods for reverse engineering of myeloid-specific regulatory networks using single-cell transcriptomics allowed us to define the key transcriptional regulators in IgAV. To further differentiate regulatory disease drivers from reactive changes, we prioritized differentially active regulators that were also co-modulated by the transcripts supported by genetic evidence. Among the 50 most differentially activated and 50 most inactivated regulators, 28 were significantly co-modulated by *FCAR, INPP5D*, and *IL6R*; moreover, their targets were significantly enriched in key immune-related pathways. These regulators appear to drive the disease process, and thus may represent attractive therapeutic targets.

Lastly, our cross-phenotype GWAS highlights shared genetic susceptibility between IgAV and IgAN, including strong genome-wide genetic correlation and shared *HLA-DRB1* risk and protective haplotypes. Most of the known IgAN-risk loci were associated with higher risk of IgAV, including the *TNFSF13* locus encoding APRIL. Phase III clinical trials of APRIL inhibition are underway for IgAN, and our results provide the first genetic support for APRIL’s involvement also in IgAV. Despite these commonalities, clear differences in the disease associations were detected at the *FCAR* and *IL6R* loci, and these differences could potentially contribute to the unique systemic manifestations of IgAV.

## Data Availability

The genetic and transcriptomic data generated by this study is being deposited in dbGAP; the accession number will be provided before publication.

## Acknowledgements

We are grateful to all study participants for their participation and contributions to this study. The study was supported by the NIH grants 1K01DK137031 (LL), 2R01DK105124 (KK, JN), 1RC2DK116690 (KK). JN and TJG are supported in part by NIH grants AI149431, DK078244, and DK082753. ATG was supported by the Ruth L. Kirschstein National Research Service Awards 5T32GM007367-43 and F30CA257765. The work of AK was supported by German Research Foundation (DFG) Project ID 431984000 (SFB 1453) and Project ID 390939984 (EXC-2189, CIBSS). The work of SMM was supported by the EQUIP Program for Medical Scientists, Faculty of Medicine, University of Freiburg. AC was supported by the NCI Outstanding Investigator award R35CA197745. The GIGA-kids Study was additionally supported by the Pediatric Nephrology Research Consortium (PNRC) and a gift from the IgA Nephropathy Foundation.

## Author contributions

All authors have read and approved the final version of manuscript. K.K. and H.Z. conceived the study, provided overall supervision of the project and made the decision to publish the findings. L.L. and L.Z. performed quality control, imputation and association analyses for GWAS discovery cohorts. J.M. and M.A.G-G. provided GWAS summary statistics for the Spanish cohort. L.L. performed GWAS meta-analysis, fine-mapping studies, functional annotations, and e/sQTL mapping. S.M. and A.K. performed transcriptome-wide and proteome-wide association studies. J.N. and T.J.G. performed structural modeling of risk variants. P.L.D. generated single cell RNA-seq datasets and L.L. and M.F. performed their processing and quality control analyses. L.L., A.T.G., L.V., and A.C. performed single cell network-based master regulator and MINDy analyses. C.B. and J.Y.Z. contributed to sample and data management. All other authors were involved in the recruitment of patients for this study.

## METHODS

We performed a GWAS meta-analysis of three large case-control cohorts comprising a total of 8,098 individuals (2,170 IgAV cases and 5,928 controls). The study cohorts are summarized in **Table S1** with detailed descriptions as follows:

### Beijing Cohort

The cases and controls were recruited from the clinics of Peking University First Hospital, Beijing, China. We used standard diagnostic criteria for IgAV that included a palpable purpura in association with at least one of the following: diffuse abdominal pain, arthritis or arthralgia, renal involvement (hematuria and/or proteinuria), and/or IgA deposition in a biopsy specimen (skin, intestinal tract, or kidney). The controls had no history of purpura or kidney disease. All participants provided informed consent to participate in genetic studies. Genomic DNA was extracted from whole blood samples using standard procedures and genotyping was performed with Illumina Omni 2.5, Omni 2.5 Exome, and Omni ZhongHua chips. The analysis of intensity clusters and genotype calls were performed in Illumina Genome Studio software 2.0. All SNPs were called on forward DNA strand and the overlapping SNPs across the three platforms were extracted for further analysis. The standard quality control (QC) included per-SNP genotyping rate >95%, per-individual genotyping rate >90%, MAF>0.01, and HWE test P>1×10^−5^ in controls. We excluded duplicates and cryptically related individuals based on the estimated pairwise kinship coefficients >0.05 using KING software^61^. Gender of each sample was imputed based on the analysis of sex chromosome markers using PLINK v1.9^62^ and individuals with gender mismatch against study records were excluded. The ancestry was evaluated using principal component analyses (PCA)^12^ using 46,494 high-quality (genotype rate > 99%), common (MAF>1%), and independent (pairwise r2<0.05) markers. The regions of high LD were excluded from this analysis (see below). Ancestry outliers were excluded based on visual inspection of PCA plots. The final PCA produced only two significant PCs by Tracy-Widom test and well-matched distribution of cases and controls (**Supplementary Figure 1a**). After QC, the final dataset comprised of 3,900 individuals (1,145 cases and 2,755 controls) genotyped with 722,465 markers with an overall genotyping rate 0.99. We carried out imputation analysis with Minimac3^63^ and pre-phasing with Eagle v2.4.1^64^ using East Asian reference populations from 1000 Genomes Project (Phase 3). A total of 6,210,008 high quality common markers (imputation R^2^>0.8 and MAF>0.01) were imputed and used in downstream analyses.

### GIGA-kids (Genomics of IgA-related disorders in kids) Cohort

This cohort included a multicenter GIGA-kids Study, as well as participants recruited by the Columbia University CKD Biobank and additional cases and controls referred through the GIGA-Europe network including collaborating sites in Italy, Poland, and France. The IgAV diagnostic criteria were the same as for the Beijing cohort and included a palpable purpura in association with at least one of the following: diffuse abdominal pain, arthritis or arthralgia, renal involvement (hematuria and/or proteinuria), and IgA deposition in a biopsy specimen (skin, intestinal tract, or kidney mesangium). The controls had no history of purpura or kidney disease. All participants provided informed consent to participate in genetic studies. DNA of whole blood samples was extracted using standard procedures and genotyped using Illumina MEGA chips. The analysis of intensity clusters and genotype calls were performed using Illumina Genome Studio 2.0; all SNPs were called on forward DNA strand. Standard QC filters included per-SNP genotyping rate >95%, per-individual genotyping rate >90%, MAF >0.01, and HWE test P>1×10^−5^ in controls as implemented in PLINK v1.9^62^. We excluded duplicates and cryptically related individuals with pairwise kinship coefficients >0.05^61^. The final post-QC dataset consisted of 2,908 individuals genotyped for 515,946 markers with an average genotyping rate of 99.9%. This cohort was further divided into two non-overlapping sub-cohorts: IgAV without nephritis sub-cohort of 316 cases and 924 controls and IgAV with nephritis (IgAVN) sub-cohort of 425 cases and 1,243 controls. For ancestry principal component analysis (PCA)^12^, we used 153,078 high-quality independent (pairwise r2<0.05) SNPs. The PCA produced 7 significant PCs in each case-control sub-cohort, and the visual inspection of PC plots confirmed comparable genetic ancestry between cases and controls (**Supplementary Figure 1b and 1c**). The imputation analysis was carried out with Minimac3^63^ after pre-phasing in Eagle V2.34^64^, and using 1000 Genomes Europeans (Phase 3) population reference. A total of 7,192,202 and 7,200,332 common high-quality markers (imputation R^2^>0.8 and MAF>0.01) were used in downstream analyses of IgAV and IgAVN sub-cohorts, respectively.

### Spanish IgAV cohort

This cohort has been described in detail before^7^. Briefly, the cohort comprised of 288 cases and 1,596 controls. The participants were recruited in Spain and were of European ancestry. All participants provided informed consent to participate in genetic studies. Genomic DNA was extracted from whole blood using standard methods and genotyped using Infinium HumanCore BeadChip (Illumina, Inc). The standard QC were performed as described before, followed by imputation using the 1000 Genomes Europeans (Phase 3) population reference. The final dataset included 5,662,805 high-quality markers (imputation R^2^>0.8 and MAF>0.01). The PCA produced 6 significant PCs and visual inspection of PC plots confirmed comparable ancestry between cases and controls (**Supplementary Figure 1d**).

### Genome-wide association studies (GWAS)

Within each cohort, primary association scans were performed using logistic regression under additive coding of dosage genotypes, and with adjustment for cohort-specific significant principal components of ancestry using PLINK v1.9^62^. No substantial genomic inflation was observed in any individual cohort (lambda <1.05 consistently for each individual cohort). Subsequently, a fixed effects meta-analysis was performed to combine the results across all cohorts using METAL^65^. Genome-wide distribution of P-values was examined visually using quantile-quantile plots for the combined analysis and the genomic inflation factor was estimated at 1.02 for the overall meta-analysis (**Supplementary Figure 2**). To declare genome-wide significance, we used the generally accepted P-value threshold of 5.0×10^−8^, while signals with P<1.0×10^−6^ were considered as suggestive. For sensitivity analysis we also performed a trans-ethnic meta-analysis under a random effects model using TransMeta software^66^, but this analysis did not change our results or the number of significant loci. To detect independent signals at genome-wide significant loci, we also performed stepwise conditional analyses of each locus in each cohort using GCTA^67^, and the conditioned summary statistics were combined across cohorts using fixed effects meta-analysis^65^.

### Transcriptome-wide association studies (TWAS)

TWAS was performed following the FUSION workflow^68^ based on weights developed from the European ancestry subset of the GTEx v8 tissues that were considered relevant to IgAV (kidney cortex, liver, spleen, whole blood, small intestine-terminal ileum, and EBV-transformed lymphocyte cells). Prediction models were based on elastic net modelling^68^. Multiple testing was accounted for by a Bonferroni adjustment for 17,126 unique number of protein-coding transcripts modelled across tissues (P < 0.05/17,126 = 2.9 × 10^−6^).

### Proteome-wide association study (PWAS)

For PWAS, we adapted the FUSION workflow^68^ using plasma protein weights from the European ancestry participants of the Atherosclerosis Risk in Communities Study (ARIC) study^69^. These weights were based on plasma proteomic quantifications using Somascan V.4.1. Prediction models were based on elastic net modelling^68^. Proteome-wide significance level was defined using a Bonferroni correction for the total number of circulating proteins tested (P < 0.05/884 = 5.7×10^−5^).

### Functional annotations of significant loci

We first defined each GWAS locus by +/-400kb of the genome-wide significant index SNP. We annotated all transcripts within these intervals using hg19 to create sets of positional candidate genes for each locus. Using ANNOVAR software^70^, we annotated all variants within the region that were in LD (r^2^>0.5) with the top SNP, including all known coding, splice, and 3’UTR and 5’UTR variants. We then performed FUN-LDA analysis to estimate the posterior probability of these variants being functional across various cell types^37^. Lastly, the variants were intersected with the predicted enhancers and mapped to regulated genes using the Activity-by-Contact (ABC) model^36^. The myeloid-specific regulatory regions were confirmed by the analysis of monocyte ATAC-seq peaks before and after LPS treatment^38^. The top prioritized variants were screened against transcription factor binding motifs using JASPAR^39^. To additionally prioritize target genes, we performed locus co-localization analyses with whole blood gene expression and splicing QTLs quantified from bulk RNA-seq of 255 IgAV cases. After harmonization of effect alleles, we identified all co-localized QTLs mapping to the region of the index SNP ±400 kb using Coloc package in R^71^. For the *IL6R* locus supported by PWAS, we additionally performed co-localization with pQTL summary statistics from participants of the ARIC study^69^. Because the pQTL signal exhibited seven conditionally independent signals (defined by the COJO-Slct algorithm^67^), we computed conditional pQTL summary statistics by conditioning on all other independently significant SNPs using COJO-Cond^67^. Subsequently, colocalization analyses were conducted for all pairwise combinations of the GWAS and conditionally independent pQTL summary statistics using an adapted version of the Giambartolomei colocalization method as implemented in the ‘coloc.fast’ function with default parameters^71^. Any co-localization with PP4>0.8 was considered as strong evidence in support of a shared causal variant. Only one of the seven conditionally independent sIL6R cis-pQTL signals co-localized with the IL6R GWAS and eQTL loci.

### HLA imputation and testing

We performed imputation of classical HLA alleles at two- and four-digit resolution, as well as individual amino acid polymorphisms at class I (*HLA-A*, *-B*, and *-C*) and class II (*HLA-DQB1*, *-DQA1* and *-DRB1*) loci using SNP2HLA software^72^. The European and East Asian cohorts were imputed separately, using ethnicity-specific reference panels. For European reference, we used the pre-phased HLA reference dataset generated by the Type 1 Diabetes Genetics Consortium (T1DGC, 5,225 individuals)^72^. For our East Asian cohorts, we used the Pan-Asian HLA Reference Panel (268 individuals)^73^. In the association analyses, we included only common HLA alleles (MAF>0.01) that were imputed with high certainty (R^2^ > 0.8). Because frequencies of HLA alleles are known to vary by ancestry, the association testing was performed separately in European and East Asian cohorts. We used logistic regression models to test the additive effects of HLA allele dosages with adjustment for significant PCs of ancestry. For multi-allelic loci, we used the following logistic regression model:

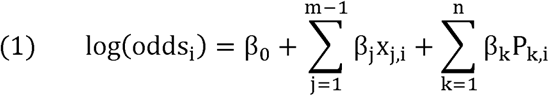

where m denotes a total number of alleles at a specific multi-allelic locus, j indicates a specific allele being tested, and x_j,i_ is the imputed dosage for allele j for individual i; β_0_ represents the intercept and β_j_ represents the additive effect of an allele j; P_k,i_ denotes the value for kt^h^ PC of individual i, n is the total number of significant PCs in the dataset, β_k_ is the effect size of principal component k. We compared log-likelihoods of the two tested models: the full model containing the test locus and relevant covariates with the reduced model without the test locus, but with the same set of covariates. The deviance was defined as -2 X log likelihood ratio, which follows a X^2^ -distribution with m - 1 degrees of freedom, from which we calculated P-values. In addition to multi-allelic tests, we also performed bi-allelic tests of association for all imputed SNPs, classical HLA alleles, and individual amino acid residues in HLA molecules. All analyses were performed using dosage method under additive coding. Stepwise conditioning analyses across the HLA region were performed using both multi-allelic and bi-allelic coding of HLA variants. In each round of stepwise conditioning, we first included the most significant variant as the covariate in the logistic regression model. If additional independently associated markers were detected, they were included as covariates in subsequent models. We repeated these analyses until no residual associations across the entire locus were observed.

### Analysis of polymorphic amino-acid sites in HLA genes

To test the effects of individual amino acid substitution sites within the three significant *HLA* genes (*HLA-DRB1, HLA-DQA1* and *HLA-DQB1*), we applied a conditional haplotype analysis using fully phased haplotypes. We tested each single amino acid position by first identifying the m possible amino-acid residues occurring at that position and then using m-1 degrees of freedom test to derive *P*-values, with a single amino acid residue arbitrarily selected as a reference. For conditioning on individual amino-acid sites, we used the following procedure: by adding a new amino acid position to the model, a total of k additional unique haplotypes were generated and tested over the null model using the likelihood ratio test with k degrees of freedom. If the new position was independently significant, we further updated the null model to include all unique haplotypes created by all amino-acid residues at both positons to identify another independent position. The procedure was repeated until no statistically significant positions were observed.

### Protein structure modeling

Models of DRα1 and DRβ1 were generated based on PDB ID: 5V4M^74^, which harbored protective residues P11 and R13. Risk variants L11/F13 and V11/H13 were generated with COOT^75^. After removing the peptide, all three models were subjected to molecular dynamics minimization protocols with YASARA^76^. Images of DRα1 and DRβ1structures were generated with ChimeraX^77^. Modeling of the IL6R-ADAM17 peptide structures followed previously published methods^46^. Briefly, ADAM17 and N-terminal residues (CTCSPSH) of the bound TIMP3 were extracted from PDB ID: 3CKI^78^. Peptide residues were mutated to 355-PVQDSSS-361 or 355-PVQASSS-361 of the IL6R stalk region, and complexes were minimized in YASARA^76^. Models of HLA-DM-HLA-DR1 were generated with PDB ID: 4GBX^79^. The parent coordinates have glutamic acid at residue 96. Variants Q96 and Y96 were generated in COOT^75^ and minimized in YASARA^76^.

### Meta-phenome-wide association study (Meta-PheWAS)

The meta-PheWAS for individual variants was performed in 786,138 participants across three biobanks: the UK Biobank (UKBB, N=460,363), Electronic Medical Records and Genomics-III (eMERGE-III, N=102,138) and All-of-Us (AoU, N=223,637). For meta-analysis, we harmonized coded diagnoses across biobanks by converting all available ICD-10 codes to the ICD-9-CM system. After conversions, we identified 1,816 unique phecodes. These phecodes were then tested using logistic regression adjusted for age, sex, study site, imputation batch, and 3 PCs of ancestry using the PheWAS R package^80^. Meta-PheWAS was performed using metagen software under fixed effects. For phenome-wide significance, we used the Bonferroni-corrected P < 2.75 × 10-5 to account for 1,816 independent phecodes tested.

### Blood RNA sequencing

Blood samples were collected using PAXGene RNA tubes from the participants of GIGA-kids study. To ensure complete lysis of blood cells and stabilization of RNA, the collected blood samples were incubated at room temperature for a minimum of 2 hours. The stabilized blood samples were further centrifuged to pellet cellular debris. Supernatant of the samples was lysed, digested, and homogenized using QuickGene RNA blood cell kit S (Catalog #: RB-S). Automated RNA extraction of lysate was done using QuickGene 810 Nucleic Acid Isolation System. RNA samples were purified with Zymo Research RNA Clean & Concentrator-5 kit (Catalog #: R1014). We use poly-A pull-down to enrich mRNAs from extracted total RNA samples with RIN>8 (200ng-1ug per sample) and performed library preparation using Illumina TruSeq RNA prep kit. Libraries were then sequenced using Illumina HiSeq2500/HiSeq4000 at Columbia University Genome Center. Sequencing generated 100bp single/pair-end reads, with a median achieved total reads per sample at 23.5M. Raw sequencing data were processed using RTA (Illumina) for base calling and bcl2fastq2 (version 2.17) for converting BCL to fastq format, coupled with adaptor trimming. RNA-seq data were aligned to the human reference genome hg38 with STAR software^81^. Following GTEx pipelines^82^, we identified and excluded low-quality samples with <10 million mapped reads, read mapping rate <0.2, intergenic mapping rate >0.3, base mismatch rate >0.01, and rRNA mapping rate >0.3. The outlier samples were further identified based on expression profile using correlation-based statistics and sex incompatibility checks following published methods^83^. After individual sample QC, we quantified the gene expression across samples using RNA-SeQC^84^ as read counts and transcripts per million (TPM). Only transcripts with TPM >0.1 and 6 read counts in at least 20% of samples were used for downstream analyses. The final post-QC dataset included 294 individuals (255 IgAV cases and 38 healthy controls) and 17,790 transcripts.

### Cell type deconvolution and differential gene expression analyses

We used CIBERSOFTx^47^ to deconvolve immune cell type abundance from bulk blood RNA-seq of 255 IgAV cases and 38 controls. We optimized this pipeline for 6 different major immune cell types, including CD4 and CD8 T cells, NK cells, monocytes, B cells and dendritic cells, using published reference panels generated by single-cell RNA-sequencing of 25,000 blood cells from 45 healthy donors^85^. The deconvolution was conducted based on the default parameters. For differential gene expression, we used DEseq2 R package^86^. This analysis was adjusted for age, sex, ancestry PCs, and deconvoluted cell-type fractions.

### Expression QTL (eQTL) mapping

For eQTL mapping, gene counts were normalized across samples using Trimmed Mean of M-values (TMM) method^87^ across 255 IgAV cases, and the expression values for each gene were inverse-normal transformed. The cis-eQTLs were identified using linear regression as implemented in fastQTL^88^, adjusting for age, sex, top 5 ancestry PCs and 45 PEER factors^89^. A false discovery rate (FDR) threshold ≤ 0.05 was applied to identify genes with at least one significant cis-eQTL (eGenes). To identify the list of all significant variant-gene pairs associated with cis-eGenes, a genome-wide empirical P-value threshold was defined as the empirical P-value of the gene closest to the 0.05 FDR threshold as descripted previously^90^. For each gene, variants with a nominal P-value below the gene-level threshold were considered significant and included in the final list of variant-gene pairs.

### Splicing QTL (sQTL) mapping

Splicing quantification was performed based on the intron excision levels across samples computed by LeafCutter^91^. Introns with few counts or with insufficient variability across samples were removed as described previously^82^. The filtered counts were normalized using the LeafCutter^91^ and the resulting splicing quantification files were used for sQTL mapping. Mapping of cis-sQTLs was performed in 255 IgAV cases using FastQTL^88^ controlling for age, sex, top 5 ancestry PCs, and 15 PEER factors^92^. To identify cis-sGenes, FDR was calculated in the same manner as for cis-eQTL, and an sGene-level nominal p-value threshold was determined for all significant variant-intron excision pairs.

### Single-cell sequencing and data processing

The reference blood single cell RNA-seq dataset was constructed using samples of volunteers recruited by the Reference Ability Neural Network (RANN) study though random mailing in Northern Manhattan^93^. Single-cell RNA-seq libraries of peripheral blood mononuclear cells (PBMC) were constructed using the 10X Genomics Chromium Next GEM Single Cell 3’ Reagent Kits as previously described^94^. An estimated 20,000 cells pooled from 3-4 donors were loaded on the 10X Genomics Chromium instrument. A total of 41 sequencing libraries (163 donors) were pooled and sequenced on the Illumina NovaSeq 6000 with S4 flow cell (Illumina, San Diego, CA) using paired-end, dual-index sequencing with 28 cycles for read 1, 10 cycles for the i7 index, 10 cycles for the i5 index, and 90 cycles for read 2. Gene expression profiles were generated for a total of 984,441 cells from 163 donors. FASTQ files were preprocessed using the count command of the CellRanger software (v6.0.0, 10x Genomics) with a human reference transcriptome GRCh38-2020-A. To de-multiplex cells from pooled samples, we took advantage of SNPs in the RNA-seq reads. Cells in the filtered Unique Molecular Identifier (UMI) count matrix were grouped into four clusters (each representing one participant) based on RNA genotypes using the freemuxlet software (downloaded on May 5^th^, 2021 from https://github.com/statgen/popscle). Cells were excluded if freemuxlet called them as doublets (N_cells_=133,117). We further excluded cells with <200 or >4500 transcripts (N_cells_=1,479), percentage mitochondrial transcripts >10% (N_cells_=44,737), and the percentage of ribosomal RNA >5% (N_cells_=12,706). We additionally removed hemoglobin genes (N_genes_=12) and genes expressed in <= 3 cells (N_genes_=681). After QC, we included 710,654 cells and 26,970 unique transcripts from 163 donors in downstream analyses. Using the Seurat R package, we performed normalization of gene expression across all 710,654 cells using LogNormalize method, followed by identification of variable transcripts, data scaling, and principal component analysis. We performed cell type clustering analysis based on the top 15 principal components. We then annotated 16 different cell types based on known blood cell markers^85^, including HSC (hematopoietic stem cells), ASC (antibody-secreting cells), Megakaryocytes, p-DC (plasmacytoid dendritic cell), c-DC (conventional dendritic cells), NK (natural killer) cells, NK CD8 T-cells, transitional or precursor cells, CD14+ CD16+ (classical) monocytes, non-classical monocytes, memory B cells, naïve B cells, effector CD4 T-cells as well as Naïve CD4 T-cells (Figure 5A).

### Myeloid-specific regulatory network construction

For reverse engineering of regulatory networks in myeloid cells, we used ARACNe3 (Algorithm for the Reconstruction of Accurate Cellular Networks version 3). This is an algorithm based on mutual information (MI) that identifies direct regulatory interactions mediated by transcription factors^48,95^. Using ARACNe3, we reconstructed a transcriptome-wide regulatory network based on myeloid cluster transcriptome generated using our reference blood single-cell RNA-seq data from 163 donors. Briefly, after QC filtering of poor-quality cells, we used 143,192 myeloid cells with UMI/cell >=3,000 across all 163 samples. We then excluded any donors with less than 500 high-quality myeloid cells, resulting in a total of 111 donors for network analysis. To address the sparsity of single cell expression matrix, we created a pseudo-bulk expression matrix by aggregating UMI counts across all myeloid cells (**Figure S11**). The ARACNe3 network was inferred using this pseudo-bulk matrix based on FDR<0.05. A total of 7 iterations were performed by random selecting 63.2% of all samples to achieve >50 targets per regulator. The final myeloid regulatory network involved 26,196 genes and 2,212 regulators based on the Gene Oncology (GO) annotations^95^.

### Master Regulator (MR) analysis

We used NaRnEA (Nonparametric analytical Rank-based Enrichment Analysis)^48^ to define differential activity of regulators based on the ARACNe-3 inferred myeloid regulons and differential gene expression signatures. NaRnEA is a gene set enrichment method which leverages a null model derived under the principle of maximum entropy. For each transcription factor, NaRnEA tests for significant enrichment of its ARACNe3-infered regulons in differential gene expression signature compared to randomly selected non-overlapping gene sets. This is important, as master regulators may be differentially active but not necessarily differentially expressed. The direction of effect was derived for each regulator (activated or suppressed in IgAV cases compared to controls), and the statistical significance was defined based on FDR<0.05. The top 50 most significant activated regulators in cases were defined as “IgAV Master Regulators”.

### Modulation of Master Regulators by candidate genes

To test how our GWAS candidate genes (*INPP5D*, *FCAR*, and *IL6R*) co-modulate IgAV Master Regulators, we performed MINDy analysis based on the myeloid regulons. MINDy is an algorithm designed to uncover modulator genes influencing transcriptional interactions. This is accomplished by the analysis of ΔMI (the difference in mutual information between a TF and its target genes) after conditioning on the highest and lowest expression of any candidate modulator. The statistical significance of a particular ΔMI for a given TF-target gene pair and a given modulator was determined based on FDR < 0.05 as descripted previously^96^.

