## Supplementary information for "Genome-wide studies define new genetic mechanisms of IgA vasculitis"

**IgA vasculitis**

*Lili Liu et al.*

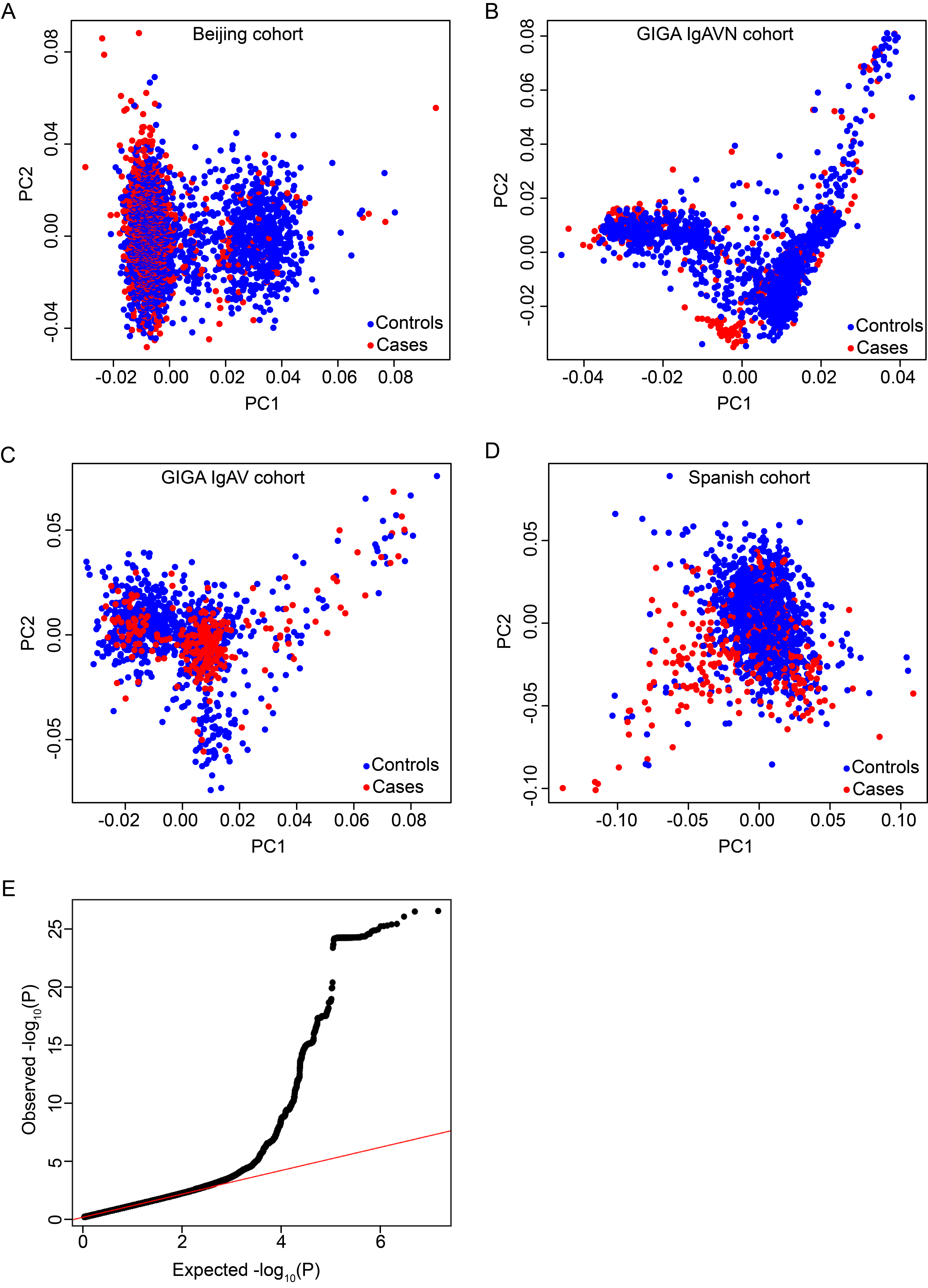

**Supplementary Figure 1. Principal component analysis (PCA) and quantile-quantile (QQ) plots. (A)** Beijing cohort of 1145 cases (red) and 2755 controls (blue). **(B)** GIGA-kids sub-cohort of 425 IgAVN cases (red) and 1243 controls (blue). **(C)** GIGA-kids IgAV sub-cohort of 315 cases (red) and 924 controls (blue). **(D)** Spanish cohort of 285 cases (red) and 1006 controls (blue). **(E)** Quantile-quantile (QQ) plot for the combined meta-analysis demonstrating no appreciable genomic inflation (lambda=1.02).

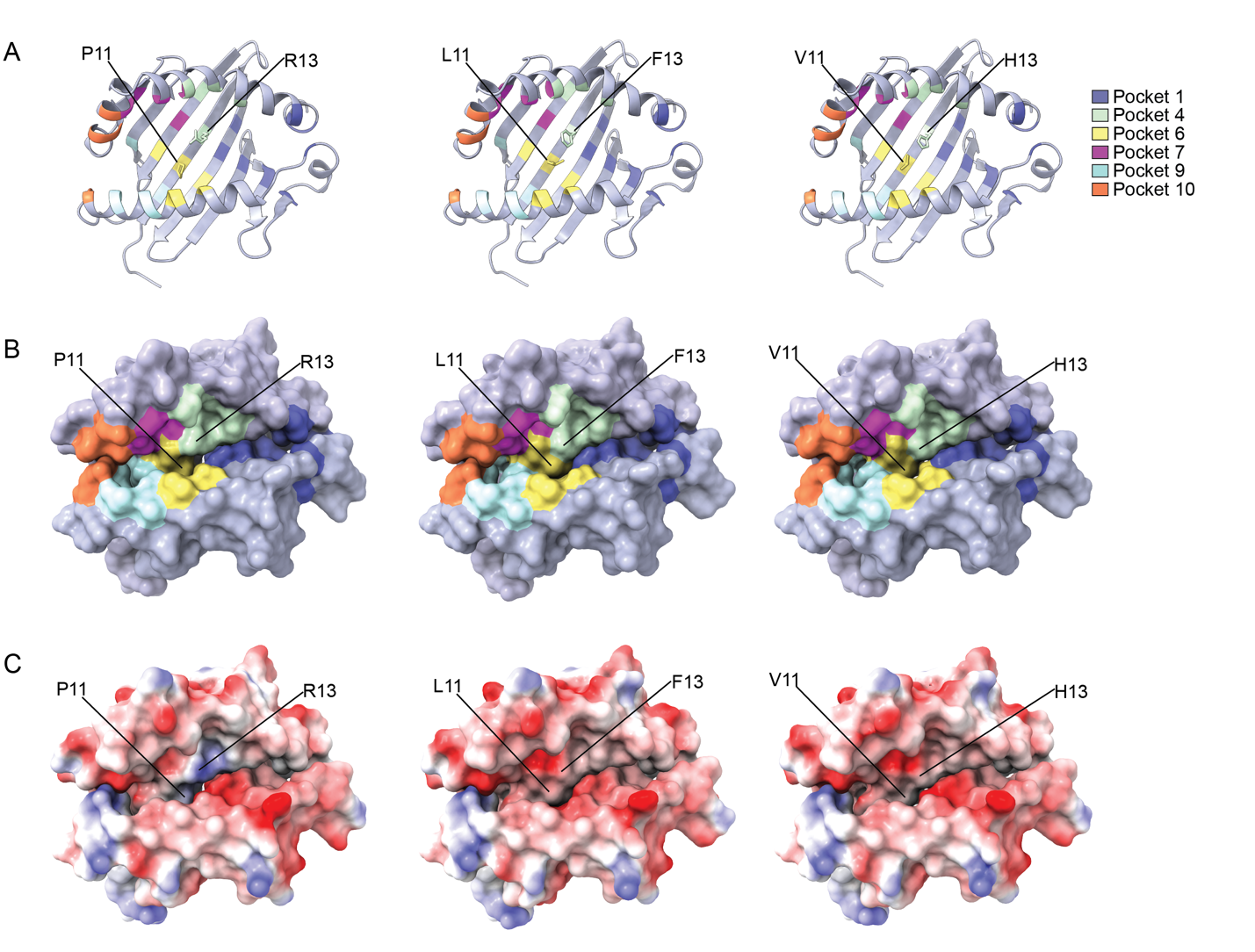
**Supplementary Figure 2.** **Structural models of DRβ1 IgAV-associated variants.** In **(A)**, protective variant (P11/R13, left) and risk variants (L11/F13, middle and V11/H13, right) are shown in ribbon model viewed into the peptide-binding groove; the residues at positions 11 and 13 of DRβ1 are shown as sticks. Surface representations are shown in **(B)** and **(C)**. In **(A)** and **(B)**, models are colored in a background of light-blue shade, with individual peptide amino-acid binding pockets shaded according to the key in **(A)**. The surface in **(C)** is colored according to electrostatic potential. All structures are based on PDB ID: 5V4M. Variants were generated with COOT. All structures with the peptide removed were subjected to molecular dynamics minimization with YASARA. Images were generated with ChimeraX.

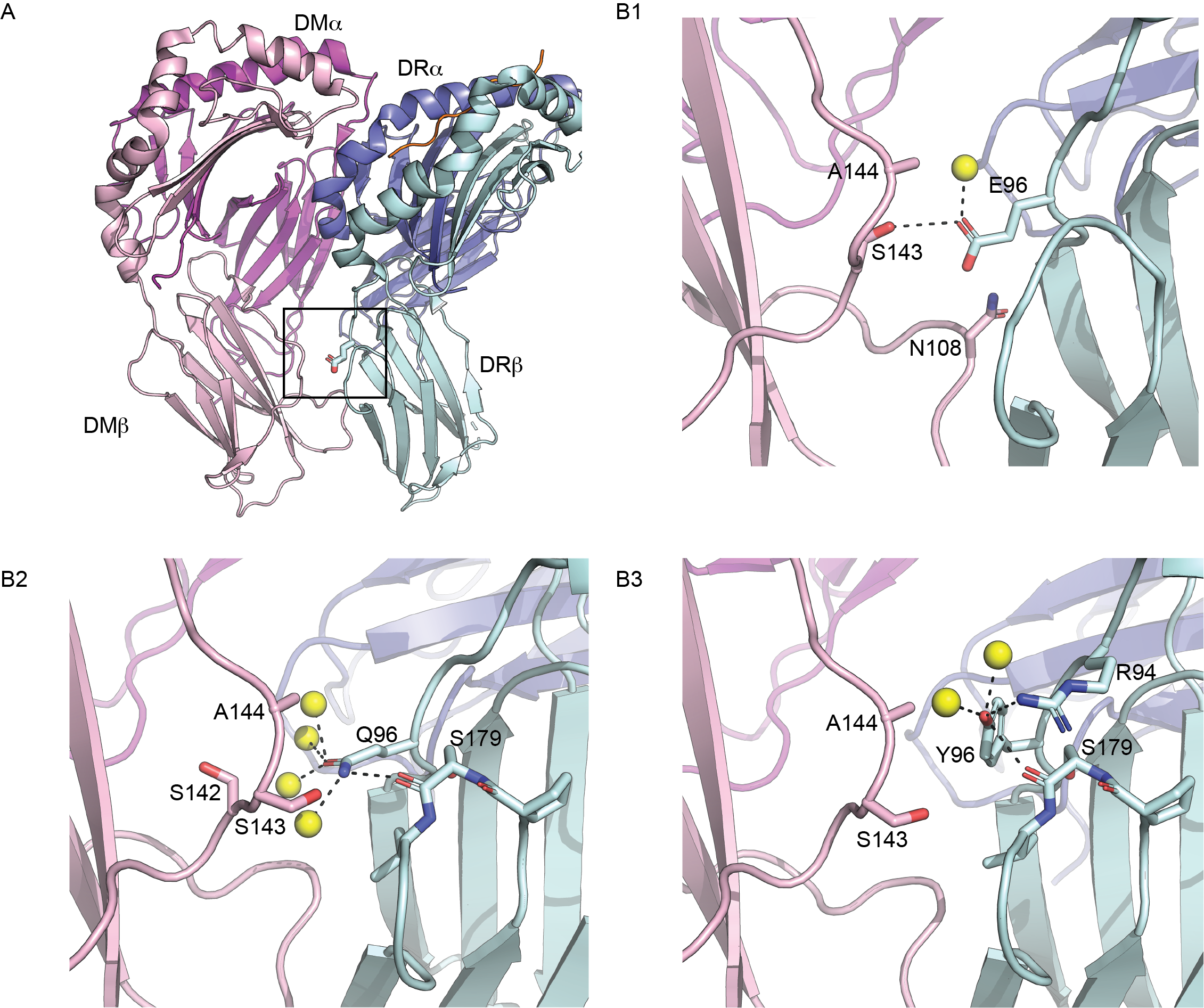

**Supplementary Figure 3. Structure of the HLA-DM-HLA-DR Complex.** **(A)** The HLA-DM–HLA-DR1 complex is shown in ribbon form, with the DM α-chain (DMα), DM β-chain (DMβ), DR1 α-chain (DRα), DR1 β-chain (DRβ) and peptide shaded in magenta, light pink, slate, pale cyan, and orange, respectively. Residue 96 of the DRβ-chain is shown in stick model. **(B1-3)** shows a close-up view of the interactions of residue 96. Amino acids within 5 angstroms of residue 96 are shown as sticks. Waters are shown as yellow spheres. Polar interactions are noted by dashed lines. **(B1 and B3)** show interactions with risk variants E96 and Y96. **(B2)** shows interactions surrounding protective variant Q96. Models in A and B1 were generated with PDB ID: 4GBX (PMID: 23260142). Variants in B2 and B3 were mutated in COOT and subjected to molecular dynamics minimization routines in YASARA. Images were generated with PyMOL.

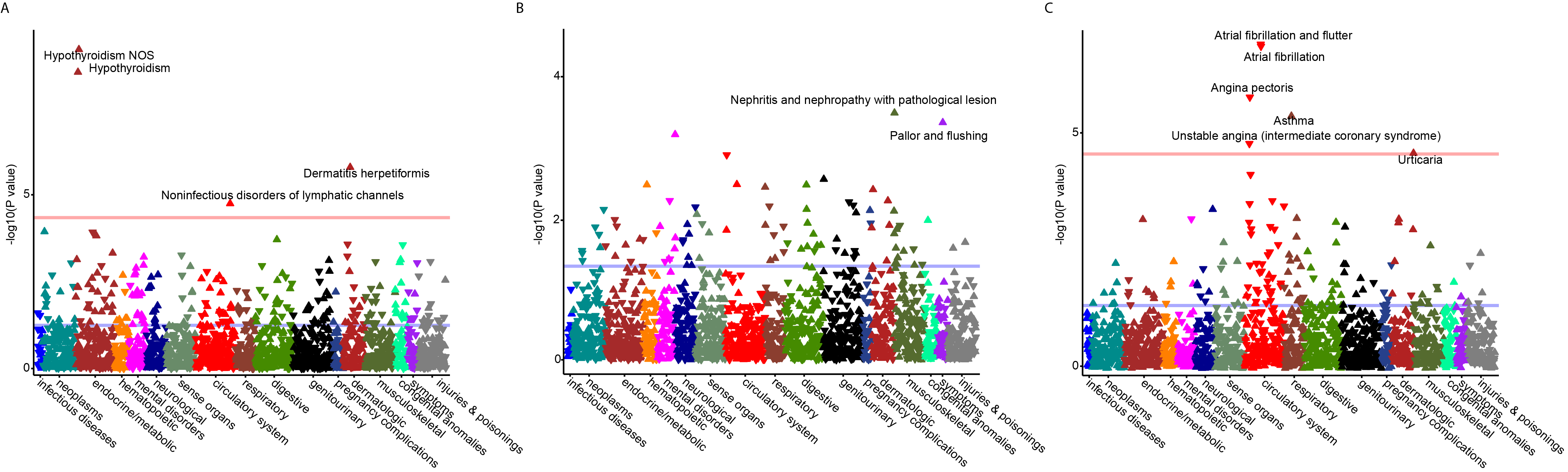

**Supplementary Figure 4. Meta-PheWAS for the top non-HLA GWAS loci across three major biobanks. (A)** the *INPP5D* locus (rs9247); **(B)** the *FCAR* locus (rs73065470); and **(C)** the *IL6R* locus (rs2228145). The meta-pheWAS was performed across the UK Biobank (UKBB, N=460,363), Electronic Medical Records and Genomics-III (eMERGE-III, N=102,138) and All of Us (AoU, N=223,637) datasets. X-axis: individual phecodes (disease codes) grouped by organ systems; Y-axis: -log of the P-value for variant association test statistic; red horizontal line indicates phenome-wide significance threshold; blue horizontal line represents a nominal statistical significance (P<0.05); an upward-pointing triangle indicates a concordant (risk) effect estimate with IgAV while a downward-pointing triangle indicates a discordant effect estimate with IgAV.

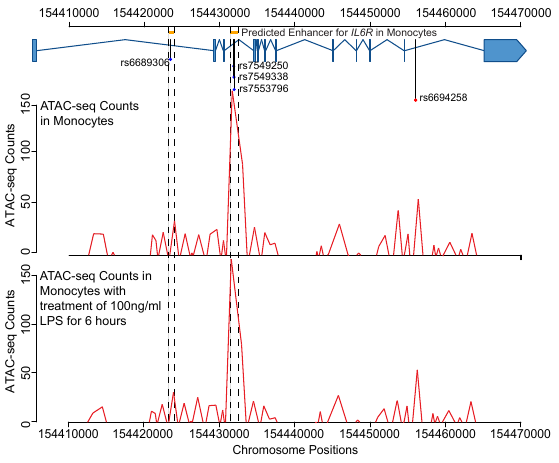

**Supplementary Figure 5. Regulatory annotations of risk alleles at the *IL6R* locus.** The upper panel shows the gene model of *IL6R*. The top SNP (rs6694258) is highlighted in red color, and four high LD SNPs with the top SNP (r2>=0.8) in blue color intersected two ABC-model predicted monocyte-specific enhancer regions (highlighted in orange segments). These regions were further confirmed by the ATAC-seq peaks in monocytes before and after treatment of 100 ng/ml LPS for 6 hours. The x-axis indicates chromosome positions and the y-axis shows the raw counts of ATAC-seq data.

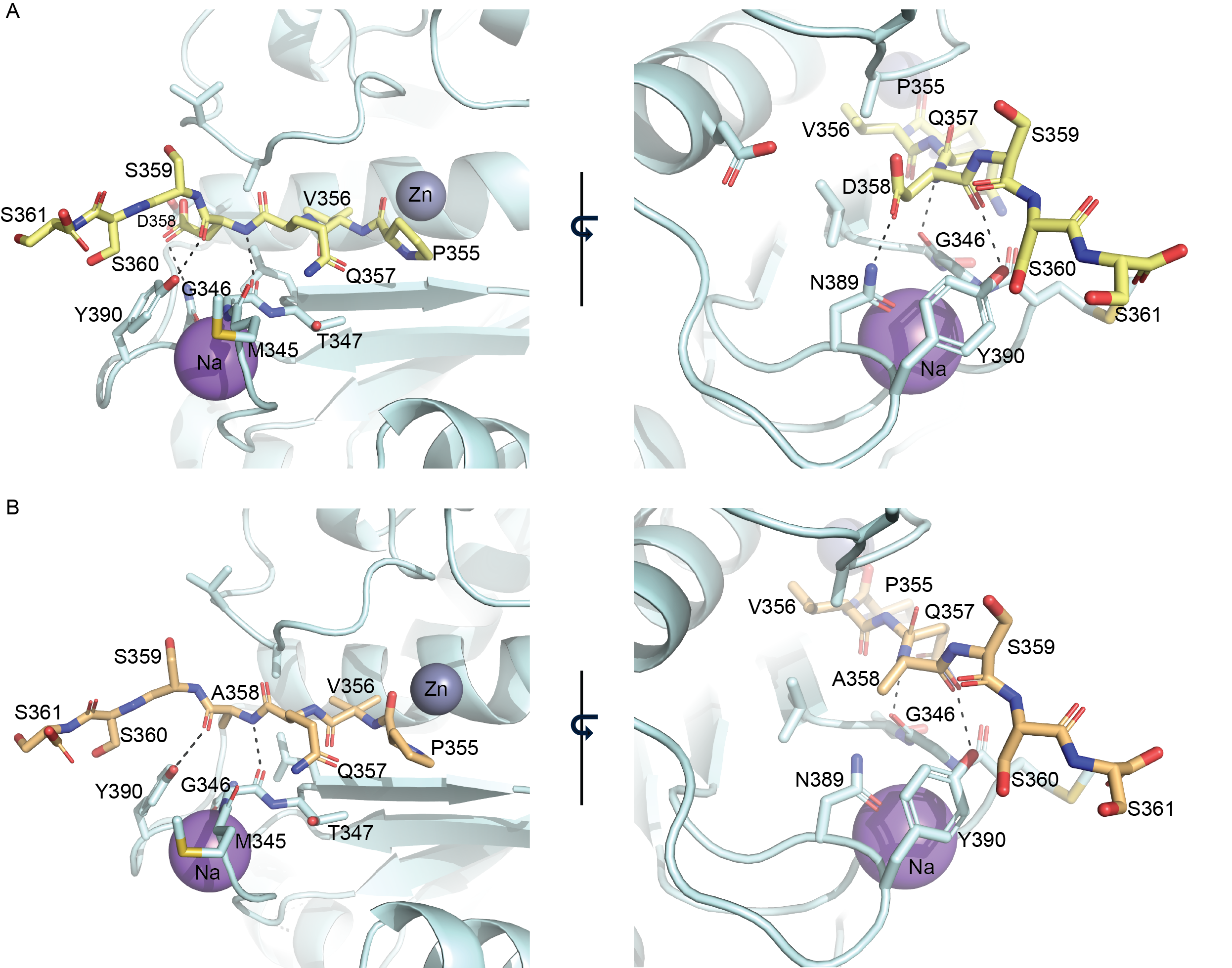

**Supplementary Figure 6.** Structure of the ADAM17 catalytic domain/IL-6R complex. The structure of ADAM17 (pale cyan ribbon render) was modeled with bound peptide 355-PVQDSSS-361 [**(A)** yellow sticks] or 355-PVQASSS-361 [**(B)** pale orange sticks] corresponding to a segment of the stalk region of IL-6R including the cleavage site (P355/V356). Residues of ADAM17 within 5 angstroms of D358 (A) or D358A (B) are shown as sticks. Polar interactions between residue 358 and ADAM17 are noted by dashed lines. The variant D358A leads to a loss of the interaction with N389 of ADAM17, potentially reducing the affinity of ADAM17 for IL-6R. Zinc and sodium ions are shown as slate and purple spheres, respectively. Images were generated with PyMOL.

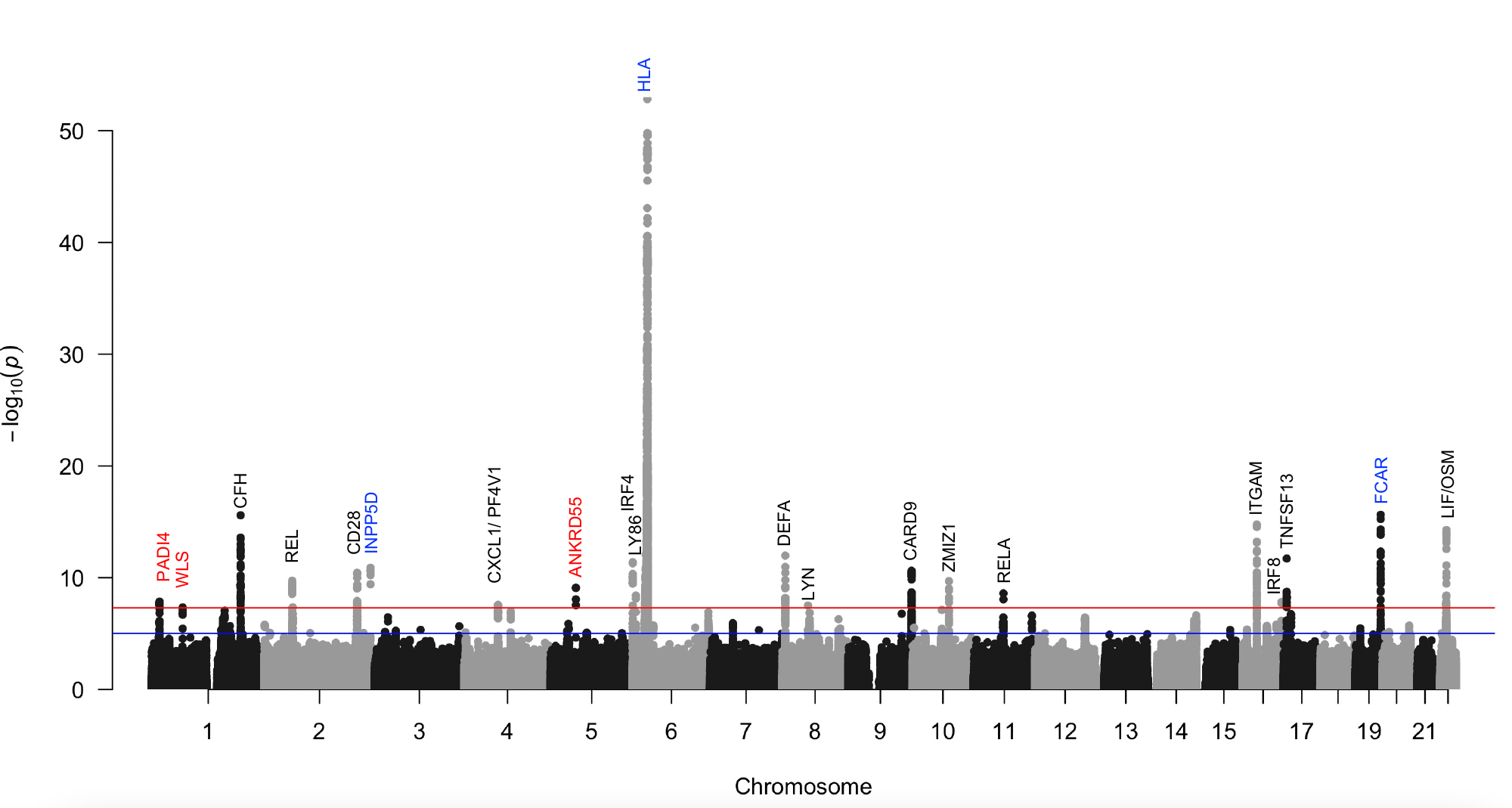

**Supplementary Figure 7. Cross-phenotype GWAS between IgAV and IgAN** identified 21 genome-wide significant loci, including 3 novel ones (labeled in red) and increased statistical significance of the IgAV loci reported in this study (labeled in blue). The red horizontal line indicates a genome-wide significance threshold (α=5×10^−8^).

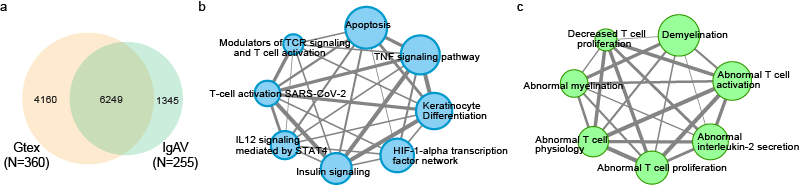

**Supplementary Figure 8. Whole blood cis-eQTL mapping in 255 IgAV cases**. (**a)** A total of 7,594 eGenes were identified in 255 IgAV cases, 1,345 eGenes were IgAV-specific compared to GTEx whole blood samples. **(b)** These IgAV-specific eGenes were significantly enriched in multiple immune-related pathways including the TNF signaling pathway and **(c)** mouse phenotypes including abnormal interleukin secretion and abnormal T cell activation. Gene set enrichment analyses in b and c were depicted as an enrichment map. Each node represents a significantly enriched pathway, the size of each node is proportional to the statistical significance of the enrichment test, each edge represents overlapping gene sets with edge thickness proportional to the number of overlapping genes (graphed in Cytoscape).

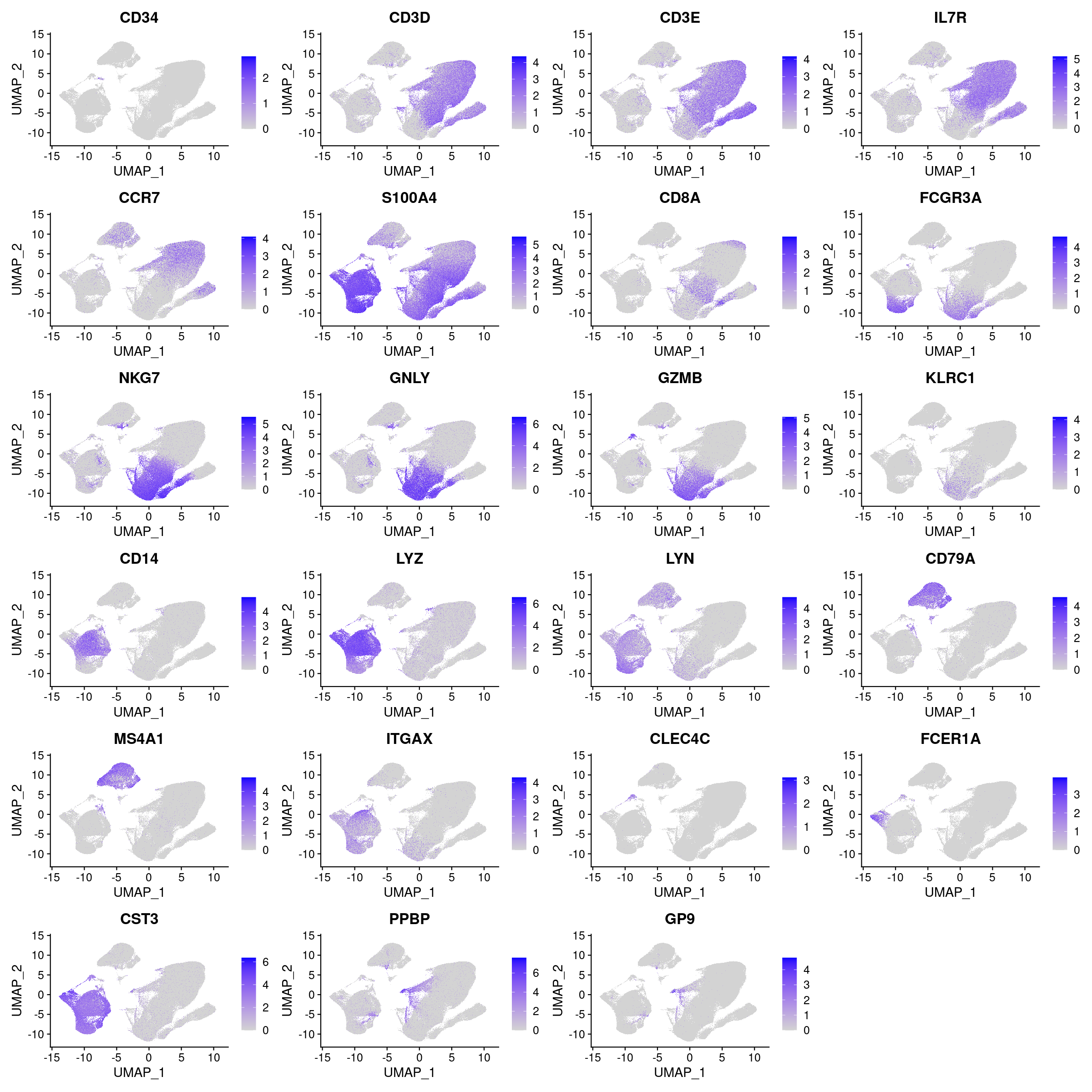

**Supplementary Figure 9. Marker expression across blood cells clusters on UMAP plots.** The x-axis denotes the values of UMAP_1, and the y-axis shows the values of UMAP_2 across cells. Colors from gray to blue indicate gene expression from low to high.

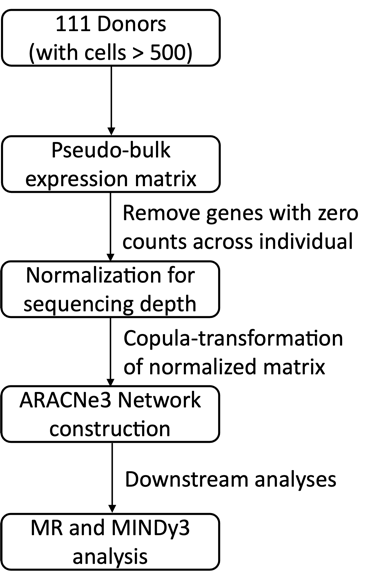

**Supplementary Figure 10. Workflow for the regulatory network construction using ARACNe3.** The network was constructed based on 111 donors with >500 high-quality myeloid cells. Next, we aggregated UMI counts across myeloid cells for each individual to generate a pseudo-bulk expression matrix as an input for network construction. The pseudo-bulk expression matrix was normalized for sequencing depth, underwent copula-transformation, and was used as input for the ARACNe3 network construction with default parameters.

**Supplementary Table 1.** Summary of study cohorts and genotyping/sequencing platforms.

| **Cohort** | **Ancestry** | **No. cases** | **No. controls** | **Total** | **Genotyping Platform** |
| --- | --- | --- | --- | --- | --- |
| **GWAS Discovery** | | | | | |
| Beijing | East Asian | 1145 | 2755 | 3900 | Omni 2.5, 2.5 Exome, ZhongHua (Illumina) |
| GIGA | European | 740 | 2167 | 2907 | MEGA chip (Illumina) |
| Spanish | European | 285 | 1006 | 1291 | HumanCore (Illumina) |
| **All combined** |  | **2170** | **5928** | **8098** |  |
| **Whole Blood RNA-seq** | | | | | |
| Columbia | Multi-ethnic | 255 | 38 | 293 | Illumina NovaSeq 6000 |
| Gtex | Multi-ethnic | 0 | 355 | 355 | Illumina NovaSeq 6000 |
| **All combined** |  | **255** | **393** | **648** |  |

**Supplementary Table 2.** Effect estimates for independent SNPs associated with IgAV risk. The trans-ethnic meta-analyses were performed under both fixed-effects and random-effects model. The stepwise conditional analysis was performed using GCTA-COJO based on combined meta-analysis results.

| **Locus** | **SNP** | **RA** | **East Asian Freq.** | **European Freq.** | **Asian Ancestry Cohorts** | | **European Ancestry Cohorts** | | **Fixed-effect META** | | **Random-effect META** | | **Q** | **I^2^** |
| --- | --- | --- | --- | --- | --- | --- | --- | --- | --- | --- | --- | --- | --- | --- |
|  |  |  |  |  | **OR  (95% CI)** | **P-value** | **OR  (95% CI)** | **P-value** | **OR  (95% CI)** | **P-value** | **OR  (95% CI)** | **P-value** |  |  |
| *INPP5D* | rs7346 | A | 0.35 | 0.19 | 1.35  (1.21-1.49) | 2.6E-08 | 1.31  (1.03-1.67) | 2.8E-02 | 1.34 (1.22-1.48) | 2.2E-09 | 1.34  (1.22-1.48) | 3.2E-09 | 0.8 | 0.0 |
| *HLA* | rs9275329 | T | 0.28 | 0.31 | 1.61  (1.44-1.81) | 2.7E-16 | 1.49  (1,32-1.67) | 3.9E-11 | 1.55 (1.43-1.68) | 1.1E-25 | 1.51  (1.30-1.75) | 1.7E-25 | 0.0 | 86.3 |
|  | rs9268791 | C | 0.71 | 0.65 | 1.36  (1.21-1.52) | 3.2E-07 | 1.58  (1.40-1.78) | 7.4E-14 | 1.46 (1.35-1.58) | 6.3E-19 | 1.48  (1.31-1.69) | 1.5E-18 | 0.9 | 0.0 |
| *FCAR* | rs77149320 | C | 0.32 | 0.26 | 1.60  (1.43-1.79) | 1.4E-16 | 1.38  (1.21-1.59) | 3.8E-06 | 1.51 (1.38-1.64) | 1.0E-20 | 1.47  (1.28-1.69) | 2.2E-20 | 0.1 | 72.5 |

| **SNP** | **CHR** | **Position** | **OR** | **P-value** | **Gene annotation** |
| --- | --- | --- | --- | --- | --- |
| rs7338986 | 13 | 71102829 | 1.25 | 3.81E-07 | *[KLHL1]* |
| rs74583145 | 17 | 7445333 | 0.72 | 4.13E-07 | *[TNFSF12-TNFSF13]* |
| rs1924200 | 13 | 38662303 | 1.22 | 1.26E-06 | *[LINC00571]* |
| rs28682450 | 15 | 91947323 | 0.62 | 1.40E-06 | *[SV2B]* |
| rs6694258 | 1 | 154428505 | 1.21 | 1.49E-06 | *[IL6R]* |
| rs2240247 | 16 | 74765354 | 0.81 | 2.76E-06 | *[FA2H, MLKL]* |
| rs475916 | 13 | 28205077 | 1.49 | 5.87E-06 | *[LNX2, POLR1D]* |
| rs6478581 | 9 | 100924221 | 0.82 | 6.43E-06 | *[CORO2A]* |
| rs7824406 | 8 | 18517026 | 0.79 | 7.09E-06 | *[PSD3]* |
| rs116695713 | 5 | 106333730 | 2.45 | 7.17E-06 | *[LOC102467213]* |
| rs1123770 | 4 | 182116420 | 0.84 | 7.65E-06 | *[intergenic]* |
| rs187231507 | 21 | 27731886 | 0.57 | 8.36E-06 | *[intergenic]* |
| rs168962 | 14 | 69282711 | 0.83 | 8.43E-06 | *[ZFP36L1]* |
| rs4875570 | 8 | 5328896 | 1.19 | 9.10E-06 | *[intergenic]* |

**Supplementary Table 3. Suggestive loci in the IgAV meta-analysis based on P-value <1x10^-5^.** P-values are based on two-sided fixed-effects meta-analysis.

| **SNP** | **Position** | **Unconditional** | | | **Conditional on *HLA-DRB1*** | | |
| --- | --- | --- | --- | --- | --- | --- | --- |
|  |  | **X2** | **df** | **p** | **X2** | **df** | **p** |
| rs660895 | 32685358 | 73.676 | 1 | 9.21E-18 | 10.641 | 1 | 1.11E-03 |
| rs9275312 | 32773706 | 72.619 | 1 | 1.57E-17 | 5.422 | 1 | 1.99E-02 |
| rs9275328 | 32774800 | 72.619 | 1 | 1.57E-17 | 5.422 | 1 | 1.99E-02 |
| rs9275374 | 32776504 | 70.939 | 1 | 3.68E-17 | 1.725 | 1 | 1.89E-01 |
| rs9275388 | 32777062 | 70.939 | 1 | 3.68E-17 | 1.725 | 1 | 1.89E-01 |
| rs9275390 | 32777134 | 70.939 | 1 | 3.68E-17 | 1.725 | 1 | 1.89E-01 |
| rs9275393 | 32777417 | 70.939 | 1 | 3.68E-17 | 1.725 | 1 | 1.89E-01 |
| rs9275406 | 32777933 | 70.939 | 1 | 3.68E-17 | 1.725 | 1 | 1.89E-01 |
| rs9275407 | 32778015 | 70.939 | 1 | 3.68E-17 | 1.725 | 1 | 1.89E-01 |
| rs9275418 | 32778222 | 70.939 | 1 | 3.68E-17 | 1.725 | 1 | 1.89E-01 |
| rs9275424 | 32778554 | 70.939 | 1 | 3.68E-17 | 1.725 | 1 | 1.89E-01 |
| rs9275427 | 32778893 | 70.939 | 1 | 3.68E-17 | 1.725 | 1 | 1.89E-01 |
| rs9275428 | 32778956 | 70.939 | 1 | 3.68E-17 | 1.725 | 1 | 1.89E-01 |
| rs9275439 | 32779499 | 70.939 | 1 | 3.68E-17 | 1.725 | 1 | 1.89E-01 |
| rs9275425 | 32778852 | 70.665 | 1 | 4.23E-17 | 1.652 | 1 | 1.99E-01 |
| AA_DRB1_13_32660109 | 32660109 | 81.450 | 4 | 8.59E-17 | 18.175 | 4 | 1.14E-03 |
| HLA_DRB1 | 32660042 | 102.919 | 12 | 1.49E-16 |  |  |  |
| AA_DRB1_37_32660037 | 32660037 | 62.647 | 4 | 8.05E-13 | 9.790 | 4 | 4.41E-02 |
| AA_DRB1_140_32657458_T | 32657458 | 51.009 | 1 | 9.20E-13 | 1.187 | 1 | 2.76E-01 |
| AA_DRB1_9_32660121 | 32660121 | 53.195 | 2 | 2.81E-12 | 2.908 | 2 | 2.34E-01 |
| HLA_DQA1 | 32716284 | 64.555 | 6 | 5.32E-12 | 19.287 | 6 | 3.70E-03 |
| AA_DRB1_120_32657518_S | 32657518 | 45.789 | 1 | 1.32E-11 | 13.786 | 1 | 2.05E-04 |
| AA_DRB1_-24_32665481_L | 32665481 | 39.782 | 1 | 2.84E-10 | 18.116 | 1 | 2.08E-05 |
| AA_DRB1_33_32660049 | 32660049 | 39.448 | 1 | 3.37E-10 | 17.915 | 1 | 2.31E-05 |
| AA_DRB1_180_32657338_V | 32657338 | 36.717 | 1 | 1.37E-09 | 13.404 | 1 | 2.51E-04 |
| AA_DRB1_96_32657590 | 32657590 | 42.701 | 3 | 2.85E-09 | 14.486 | 3 | 2.31E-03 |
| HLA_DQB1 | 32739039 | 45.595 | 4 | 2.99E-09 | 30.961 | 12 | 2.00E-03 |
| AA_DQB1_140_32737868_T | 32737868 | 33.683 | 1 | 6.48E-09 | 2.807 | 1 | 9.39E-02 |
| AA_DQB1_182_32737742_S | 32737742 | 33.529 | 1 | 7.02E-09 | 2.722 | 1 | 9.90E-02 |
| AA_DQA1_52_32717206 | 32717206 | 37.077 | 2 | 8.89E-09 | 6.138 | 2 | 4.65E-02 |
| AA_DQA1_69_32717257 | 32717257 | 36.739 | 2 | 1.05E-08 | 8.600 | 2 | 1.36E-02 |
| AA_DQA1_47_32717191 | 32717191 | 38.696 | 3 | 2.01E-08 | 15.114 | 3 | 1.72E-03 |

**Supplementary Table 4. Stepwise conditioning analysis across HLA region based on multi-allelic coding of the HLA variants in East Asians.** This table includes multi-allelic variants with unconditional P-value < 5x10^-8^. After conditioning on the most significant HLA gene (*HLA-DRB1*), there is no additional genome-wide significant residual. df: The multi-degree of freedom; χ2: test statistics; p: p-values.

**Supplementary Table 5. Stepwise conditional analysis of single amino-acid sites within *HLA-DRB1* (A), -*DQA1* (B) and -*DQB1* (C) in East Asians:** multi-degree of freedom (df) test statistics χ2 and p-values.

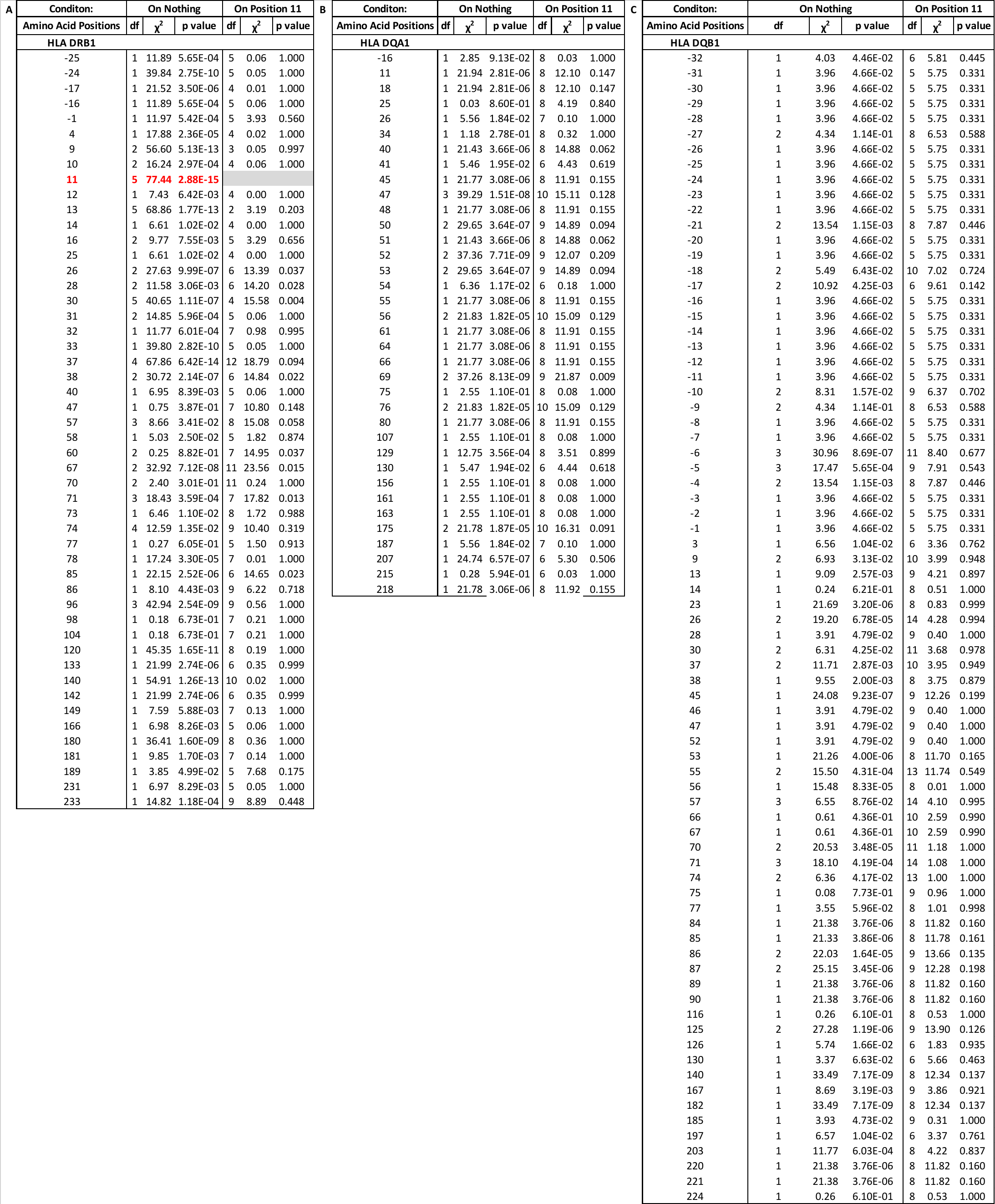

**Supplementary Table 6. Effects of individual amino-acid residues at position 11 of the *HLA-DRB1* gene in East Asian and European ancestry cohorts separately.** FRQcase indicates the allele frequency in cases; FRQctrl shows the allelic frequency in controls; OR: Odds Ratio; SE: Standard Error; P: P-value.

| **Amino acid substitution (DRB1 position 11)** | **East Asian** | | | | | **European** | | | | |
| --- | --- | --- | --- | --- | --- | --- | --- | --- | --- | --- |
|  | **FRQcase** | **FRQctrl** | **OR** | **SE** | **P** | **FRQcase** | **FRQctrl** | **OR** | **SE** | **P** |
| V | 0.180 | 0.121 | 1.654 | 0.072 | 6.77E-12 | 0.174 | 0.109 | 1.417 | 0.083 | 2.61E-05 |
| P | 0.164 | 0.202 | 0.734 | 0.068 | 5.48E-06 | 0.112 | 0.145 | 0.648 | 0.095 | 4.85E-06 |
| D | 0.114 | 0.152 | 0.789 | 0.079 | 0.00187 | -- | -- | -- | -- | -- |
| S | 0.417 | 0.395 | 1.151 | 0.052 | 0.00631 | 0.400 | 0.448 | 0.850 | 0.063 | 0.01 |
| G | 0.098 | 0.104 | 0.804 | 0.086 | 0.009743 | 0.105 | 0.148 | 0.831 | 0.092 | 0.043 |
| L | 0.029 | 0.029 | 0.919 | 0.150 | 0.4053 | 0.174 | 0.109 | 1.720 | 0.085 | 1.686E-10 |

**Supplementary Table 7. Stepwise conditioning analysis across the HLA region based on multi-allelic coding of variants in participants of European ancestry.** This table includes the multi-allelic variants with unconditional P-value < 5x10^-8^. After conditioning on the most significant HLA gene (*HLA DRB1*), there is no additional genome-wide significant residuals. Df: The multi-degree of freedom; χ2: test statistics; p: p-values.

| **SNP** | **Position** | **Unconditional** | | | **Conditional on HLA-DRB1** | | |
| --- | --- | --- | --- | --- | --- | --- | --- |
|  |  | **X2** | **df** | **p** | **X2** | **df** | **p** |
| SNP_DRB1_32656559 | 32656559 | 62.52 | 1 | 2.64E-15 | 0.17 | 1 | 6.79E-01 |
| SNP_DRB1_32657591 | 32657591 | 64.83 | 2 | 8.35E-15 | 0.64 | 2 | 7.27E-01 |
| rs9268557 | 32497283 | 56.95 | 1 | 4.46E-14 | 6.34 | 1 | 1.18E-02 |
| rs7764856 | 32788618 | 50.83 | 1 | 1.00E-12 | 0.05 | 1 | 8.27E-01 |
| rs7754768 | 32528157 | 50.63 | 1 | 1.12E-12 | 5.70 | 1 | 1.70E-02 |
| rs9268832 | 32535767 | 48.93 | 1 | 2.65E-12 | 4.26 | 1 | 3.91E-02 |
| rs9275582 | 32788048 | 48.47 | 1 | 3.36E-12 | 0.53 | 1 | 4.67E-01 |
| rs9275578 | 32787362 | 48.46 | 1 | 3.37E-12 | 0.53 | 1 | 4.66E-01 |
| rs13193645 | 32704302 | 48.32 | 1 | 3.61E-12 | 5.24 | 1 | 2.21E-02 |
| rs9275555 | 32785066 | 47.32 | 1 | 6.02E-12 | 0.73 | 1 | 3.94E-01 |
| HLA_DQA1 | 32716284 | 76.74 | 11 | 6.29E-12 | 10.00 | 6 | 1.24E-01 |
| rs9275580 | 32787440 | 46.76 | 1 | 8.03E-12 | 0.84 | 1 | 3.60E-01 |
| SNP_DQB1_32740689 | 32740689 | 45.55 | 1 | 1.49E-11 | 3.78 | 1 | 5.20E-02 |
| SNP_DRB1_32657589 | 32657589 | 49.84 | 2 | 1.50E-11 | 0.97 | 2 | 6.15E-01 |
| rs8084 | 32519013 | 44.64 | 1 | 2.37E-11 | 7.37 | 1 | 6.63E-03 |
| rs3998158 | 32789970 | 44.03 | 1 | 3.24E-11 | 0.45 | 1 | 5.03E-01 |
| rs3130320 | 32331236 | 43.51 | 1 | 4.22E-11 | 13.67 | 1 | 2.18E-04 |
| HLA_DQB1 | 32739039 | 82.20 | 16 | 6.65E-11 | 31.95 | 11 | 7.78E-04 |
| SNP_DQA1_32717151 | 32717151 | 42.43 | 1 | 7.34E-11 | 1.09 | 1 | 2.96E-01 |
| AA_DQA1_34_32717152 | 32717152 | 42.43 | 1 | 7.34E-11 | 1.09 | 1 | 2.96E-01 |
| SNP_DRB1_32657592 | 32657592 | 42.16 | 1 | 8.41E-11 | 0.64 | 1 | 4.25E-01 |
| AA_DRB1_26_32660070_L | 32660070 | 49.58 | 3 | 9.83E-11 | 5.37 | 3 | 1.47E-01 |
| SNP_DRB1_32660045 | 32660045 | 41.84 | 1 | 9.93E-11 | 0.96 | 1 | 3.27E-01 |
| rs4530903 | 32689867 | 41.78 | 1 | 1.02E-10 | 1.29 | 1 | 2.56E-01 |
| rs9275482 | 32780910 | 41.49 | 1 | 1.18E-10 | 0.49 | 1 | 4.86E-01 |
| rs3830135 | 32656442 | 40.70 | 1 | 1.77E-10 | 0.63 | 1 | 4.26E-01 |
| SNP_DRB1_32657421 | 32657421 | 40.70 | 1 | 1.77E-10 | 0.63 | 1 | 4.26E-01 |
| rs17533167 | 32698822 | 40.70 | 1 | 1.77E-10 | 0.63 | 1 | 4.26E-01 |
| SNP_DRB1_32660105 | 32660105 | 40.70 | 1 | 1.77E-10 | 0.63 | 1 | 4.26E-01 |
| rs9275425 | 32778852 | 40.42 | 1 | 2.05E-10 | 0.68 | 1 | 4.11E-01 |
| rs9275371 | 32776274 | 40.38 | 1 | 2.09E-10 | 0.67 | 1 | 4.13E-01 |
| rs9275332 | 32774921 | 40.37 | 1 | 2.10E-10 | 0.67 | 1 | 4.13E-01 |
| rs9275390 | 32777134 | 40.33 | 1 | 2.15E-10 | 0.68 | 1 | 4.09E-01 |
| rs9275393 | 32777417 | 40.33 | 1 | 2.15E-10 | 0.68 | 1 | 4.09E-01 |
| rs9275407 | 32778015 | 40.33 | 1 | 2.15E-10 | 0.68 | 1 | 4.09E-01 |
| rs9275424 | 32778554 | 40.33 | 1 | 2.15E-10 | 0.68 | 1 | 4.09E-01 |
| rs9275439 | 32779499 | 40.33 | 1 | 2.15E-10 | 0.68 | 1 | 4.09E-01 |
| rs9275428 | 32778956 | 40.11 | 1 | 2.41E-10 | 0.73 | 1 | 3.93E-01 |
| rs17427599 | 32775342 | 39.83 | 1 | 2.77E-10 | 1.18 | 1 | 2.77E-01 |
| HLA_DRB1 | 32660042 | 97.69 | 26 | 3.10E-10 |  |  |  |
| rs644045 | 31991936 | 39.42 | 1 | 3.43E-10 | 16.94 | 1 | 3.87E-05 |
| rs1794275 | 32779226 | 39.21 | 1 | 3.81E-10 | 4.12 | 1 | 4.24E-02 |
| rs7192 | 32519624 | 38.46 | 1 | 5.59E-10 | 6.62 | 1 | 1.01E-02 |
| rs3763327 | 32521808 | 38.36 | 1 | 5.89E-10 | 6.56 | 1 | 1.04E-02 |
| rs7195 | 32520517 | 38.31 | 1 | 6.05E-10 | 6.51 | 1 | 1.07E-02 |
| rs2213586 | 32521072 | 38.31 | 1 | 6.05E-10 | 6.51 | 1 | 1.07E-02 |
| rs2213585 | 32521128 | 38.31 | 1 | 6.05E-10 | 6.51 | 1 | 1.07E-02 |
| rs2227139 | 32521437 | 38.31 | 1 | 6.05E-10 | 6.51 | 1 | 1.07E-02 |
| rs17496549 | 32517686 | 37.78 | 1 | 7.94E-10 | 1.38 | 1 | 2.40E-01 |
| rs17533090 | 32698700 | 37.53 | 1 | 9.01E-10 | 6.06 | 1 | 1.39E-02 |
| AA_DRB1_96_32657590_Q | 32657590 | 68.75 | 13 | 1.36E-09 | 1.72 | 13 | 1.00E+00 |
| rs17496307 | 32509014 | 36.68 | 1 | 1.40E-09 | 0.96 | 1 | 3.27E-01 |
| rs3130316 | 32329206 | 35.22 | 1 | 2.94E-09 | 10.12 | 1 | 1.47E-03 |
| rs521977 | 31944806 | 35.14 | 1 | 3.07E-09 | 9.00 | 1 | 2.69E-03 |
| rs3115572 | 32328462 | 35.09 | 1 | 3.15E-09 | 10.01 | 1 | 1.56E-03 |
| rs592229 | 32038420 | 35.03 | 1 | 3.25E-09 | 14.70 | 1 | 1.26E-04 |
| rs9271588 | 32698931 | 34.51 | 1 | 4.24E-09 | 0.53 | 1 | 4.68E-01 |
| rs3129890 | 32522251 | 34.19 | 1 | 5.00E-09 | 8.53 | 1 | 3.50E-03 |
| rs6936204 | 32325070 | 33.66 | 1 | 6.55E-09 | 6.56 | 1 | 1.04E-02 |
| rs13192471 | 32779081 | 33.60 | 1 | 6.75E-09 | 1.20 | 1 | 2.73E-01 |
| rs2734335 | 32001923 | 33.57 | 1 | 6.86E-09 | 11.11 | 1 | 8.57E-04 |
| rs589428 | 31956199 | 33.54 | 1 | 7.00E-09 | 11.32 | 1 | 7.68E-04 |
| rs17499655 | 32780113 | 33.39 | 1 | 7.53E-09 | 1.10 | 1 | 2.95E-01 |
| rs535586 | 31968316 | 33.36 | 1 | 7.65E-09 | 11.34 | 1 | 7.58E-04 |
| rs204992 | 32264886 | 33.12 | 1 | 8.68E-09 | 10.99 | 1 | 9.17E-04 |
| rs176095 | 32266297 | 32.67 | 1 | 1.09E-08 | 10.69 | 1 | 1.08E-03 |
| rs204994 | 32262976 | 32.66 | 1 | 1.10E-08 | 10.49 | 1 | 1.20E-03 |
| rs1964995 | 32557389 | 32.55 | 1 | 1.16E-08 | 0.16 | 1 | 6.93E-01 |
| SNP_DRB1_32657334 | 32657334 | 32.54 | 1 | 1.17E-08 | 0.16 | 1 | 6.92E-01 |
| SNP_DRB1_32660056_C | 32660056 | 36.21 | 2 | 1.37E-08 | 0.38 | 2 | 8.27E-01 |
| AA_DRB1_31_32660055_F | 32660055 | 36.21 | 2 | 1.37E-08 | 0.38 | 2 | 8.27E-01 |
| SNP_DRB1_32660069_GT | 32660069 | 45.04 | 5 | 1.43E-08 | 4.45 | 5 | 4.86E-01 |
| SNP_DRB1_32659948_G | 32659948 | 35.50 | 2 | 1.96E-08 | 3.14 | 2 | 2.08E-01 |
| AA_DRB1_67_32659947_F | 32659947 | 35.50 | 2 | 1.96E-08 | 3.14 | 2 | 2.08E-01 |
| rs3129891 | 32523058 | 31.23 | 1 | 2.29E-08 | 8.92 | 1 | 2.82E-03 |
| SNP_DRB1_32659976 | 32659976 | 31.22 | 1 | 2.31E-08 | 0.07 | 1 | 7.98E-01 |
| SNP_DQB1_32737837 | 32737837 | 30.29 | 1 | 3.71E-08 | 0.51 | 1 | 4.74E-01 |
| rs7774434 | 32765556 | 30.06 | 1 | 4.19E-08 | 0.01 | 1 | 9.10E-01 |

**Supplementary Table 8. Stepwise conditional analysis of single amino-acid sites within *HLA-DRB1* (A), -*DQA1* (B) and -*DQB1* (C) in participants of European ancestry:** multi-degree of freedom (df) test statistics χ2 and p-values.

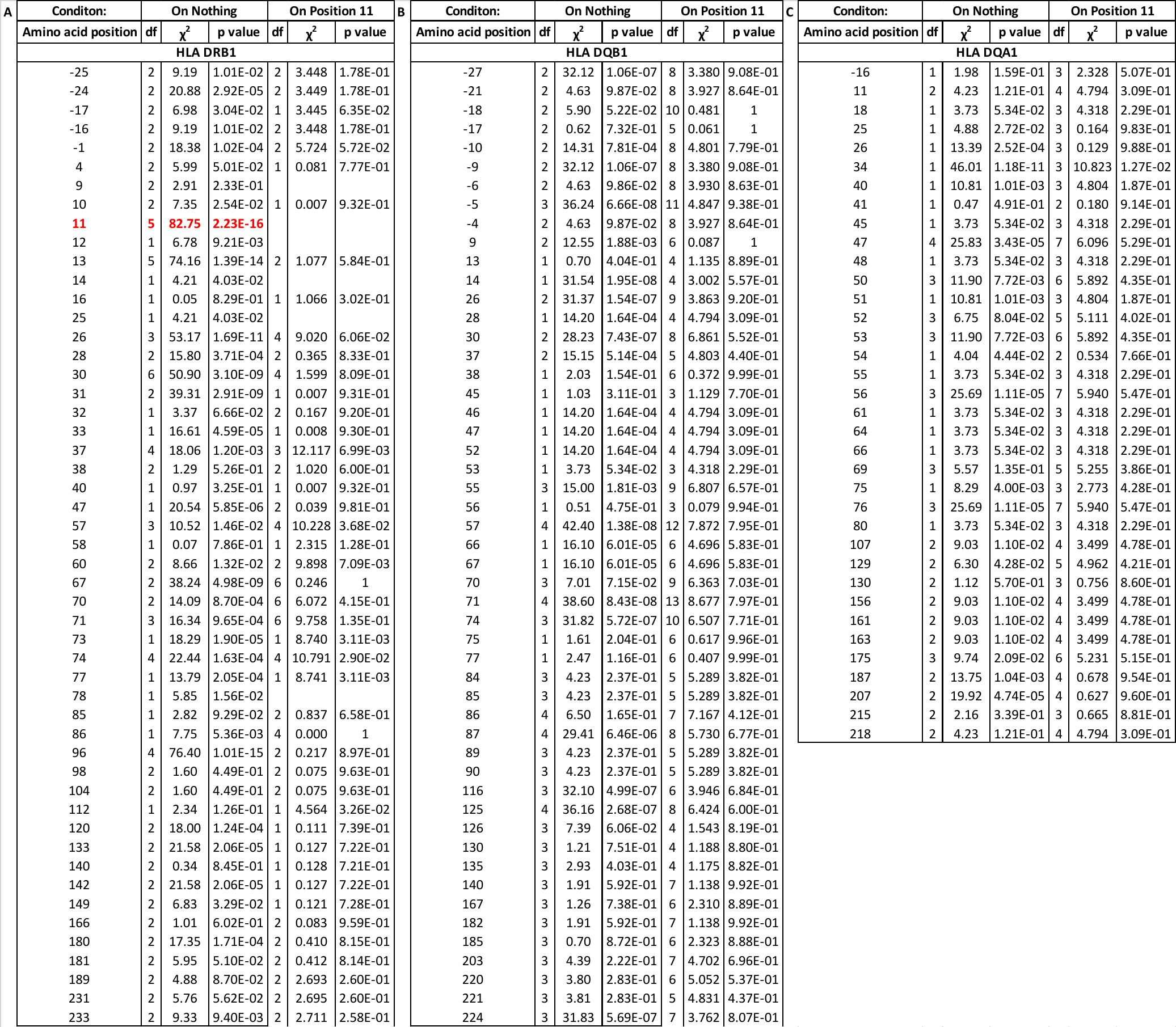

**Supplementary Table 9. ANNOVAR annotation of the top significant non-HLA SNPs and their proxies (R2 >0.5).** P-values are based on fixed effects meta-analysis. Only SNPs that have ANNOVAR annotations are included in the table.

| **Locus** | **SNP** | **Chr.** | **Position (Hg19)** | **Risk allele** | **OR** | **P (discovery)** | **R^2^ with the top SNP in GWAS** | **Annotation** |
| --- | --- | --- | --- | --- | --- | --- | --- | --- |
| INPP5D | rs7346 | 2 | 234113057 | A | 1.34 | 2.22E-09 | 1 | Synonymous variant in INPP5D (Top SNP) |
|  | rs9247 | 2 | 234113301 | T | 1.34 | 2.26E-09 | 0.99 | Missense variant (p.His1157Tyr) in INPP5D |
| FCAR | rs77149320 | 19 | 55388245 | C | 1.51 | 1.04E-20 | 1 | intronic (Top SNP) |
|  | rs3816051 | 19 | 55385604 | C | 1.28 | 7.23E-11 | 0.54 | UTR5 of FCAR |

**Supplementary Table 10. Top SNPs intersecting ABC model-predicted enhancer regions: (a)** the *FCAR* locus**; (b)** the *IL6R* locus. There was no SNP intersecting enhancer region for the *INPP5D* locus. The effect estimates and P-values are based on GWAS meta-analysis for IgAV. All listed enhancer regions were predicted based on the ABC model, including a corresponding target gene and cell type. Only SNPs with LD >0.8 with the index SNP per each locus were included.

| **SNP** | **GWAS for IgAV** | | | **Enhancer Name** | **Target Gene** | **Cell Type** | **R2 with top SNP** |
| --- | --- | --- | --- | --- | --- | --- | --- |
|  | **RA** | **OR** | **P value** |  |  |  |  |
| **(a) *FCAR* Locus** | |  |  |  |  |  |  |
| rs77149320 | C | 1.51 | 1.04E-20 | -- | -- | -- | 1 |
| rs28642682 | G | 1.43 | 2.31E-17 | genic\|chr19:55388836-55389336 | *FCAR* | CD14+ monocyte | 0.83 |
|  |  |  |  | genic\|chr19:55388872-55389372 |  | dendritic cell |  |
|  |  |  |  | genic\|chr19:55388928-55389428 |  |  |  |
|  |  |  |  | genic\|chr19:55388955-55389455 |  |  |  |
| rs28536683 | C | 1.43 | 2.472E-17 | genic\|chr19:55388836-55389336 | *FCAR* | CD14+ monocyte | 0.83 |
|  |  |  |  | genic\|chr19:55388872-55389372 |  | dendritic cell |  |
|  |  |  |  | genic\|chr19:55388928-55389428 |  |  |  |
|  |  |  |  | genic\|chr19:55388955-55389455 |  |  |  |
| rs4806602 | G | 1.43 | 1.775E-17 | genic\|chr19:55388836-55389336 | *FCAR* | CD14+ monocyte | 0.82 |
|  |  |  |  | genic\|chr19:55388872-55389372 |  | dendritic cell |  |
| **(b) *IL6R* Locus** | |  |  |  |  |  |  |
| rs6689306 | G | 1.2 | 1.24E-05 | genic\|chr1:154395707-154396428 | *IL6R* | dendritic cell | 0.90 |
| rs7549250 | T | 1.2 | 0.000007655 | genic\|chr1:154403909-154404918 | *IL6R* | dendritic cell | 0.89 |
|  |  |  |  | genic\|chr1:154403910-154404519 | *IL6R* | CD14+ monocyte |  |
|  |  |  |  | genic\|chr1:154403985-154404706 | *IL6R, ATP8B2, HAX1, SHE, TDRD10* | dendritic cell |  |
|  |  |  |  | genic\|chr1:154403988-154404488 | *IL6R* | dendritic cell |  |
|  |  |  |  | genic\|chr1:154403989-154404489 | *IL6R* | CD14+ monocyte |  |
| rs7549338 | G | 1.19 | 0.000009204 | genic\|chr1:154403909-154404918 | *IL6R* | CD14+ monocyte | 0.91 |
|  |  |  |  | genic\|chr1:154403985-154404706 | *IL6R, ATP8B2, HAX1, SHE, TDRD10* | dendritic cell |  |
| rs7553796 | C | 1.19 | 0.000009366 | genic\|chr1:154403909-154404918 | *IL6R* | CD14+ monocyte | 0.91 |
|  |  |  |  | genic\|chr1:154403985-154404706 | *IL6R, ATP8B2, HAX1, SHE, TDRD10* | dendritic cell |  |

**Supplementary Table 11. Cross-phenotype GWAS meta-analysis for IgA vasculitis (IgAV) and IgA nephropathy (IgAN) by MTAG:** 21 genome-wide significant loci including three novel ones (*PADI3/4*, *WLS,* and *ANKRD55*). The effect estimates (OR), 95% confidence intervals (95CI), P-values (P), and heterogeneity metrics between the two traits (Q: p-value for the Cochrane heterogeneity test and I^2^ heterogeneity index)

| **Locus** | **SNP** | **CHR** | **BP (hg19)** | **A1** | **A2** | **HSPN GWAS** | | | **IgAN GWAS** | | | **Cross Phenotype Meta-analysis** | | | | |
| --- | --- | --- | --- | --- | --- | --- | --- | --- | --- | --- | --- | --- | --- | --- | --- | --- |
|  |  |  |  |  |  | **OR** | **95 CI** | **P** | **OR** | **95 CI** | **P** | **OR** | **95 CI** | **P** | **Q** | **I^2^** |
| *PADI3/4* | rs2501784 | 1 | 17630193 | A | G | 1.11 | 1.03-1.20 | 5.3E-03 | 1.11 | 1.07-1.15 | 1.2E-07 | 1.05 | 1.03-1.06 | 1.4E-08 | 0.9 | 0 |
| *CFH* | rs6677604 | 1 | 196686918 | A | G | 0.89 | 0.79-0.99 | 3.4E-02 | 0.79 | 0.75-0.83 | 1.5E-17 | 0.92 | 0.90-0.94 | 2.6E-16 | 0.1 | 71 |
| *WLS^a^* | rs10493441 | 1 | 68720758 | A | G | 1.16 | 1.05-1.27 | 3.6E-03 | 1.15 | 1.09-1.22 | 6.3E-07 | 1.05 | 1.03-1.07 | 4.7E-08 | 1.0 | 0 |
| *CD28* | rs3769684 | 2 | 204584759 | T | C | 1.18 | 1.02-1.24 | 1.1E-03 | 1.17 | 1.13-1.25 | 3.1E-09 | 1.05 | 1.04-1.07 | 3.6E-11 | 1.0 | 0 |
| *INPP5D^b,c^* | rs13408498 | 2 | 234114942 | T | C | 0.75 | 0.68-0.83 | 3.7E-09 | 0.88 | 0.83-0.92 | 3.3E-07 | 0.95 | 0.93-0.96 | 1.3E-11 | 0 | 87 |
| *REL* | rs842647 | 2 | 61119471 | A | G | 0.85 | 0.77-0.93 | 4.6E-04 | 0.88 | 0.84-0.92 | 7.9E-09 | 0.95 | 0.94-0.97 | 1.9E-10 | 0.5 | 0 |
| *PF4V1* | rs7688774 | 4 | 74723372 | T | C | 0.91 | 0.84-1.00 | 4.1E-02 | 0.86 | 0.82-0.91 | 3.8E-08 | 0.95 | 0.93-0.97 | 2.7E-08 | 0.3 | 15 |
| *ANKRD55^a,c^* | rs6859219 | 5 | 55438580 | A | C | 0.71 | 0.60-0.83 | 4.2E-05 | 0.82 | 0.79-0.90 | 4.2E-06 | 0.94 | 0.93-0.96 | 8.1E-10 | 0.1 | 74 |
| *IRF4/DUSP22* | rs12201499 | 6 | 249571 | T | C | 0.83 | 0.73-0.94 | 2.6E-03 | 0.82 | 0.77-0.87 | 3.1E-11 | 0.94 | 0.92-0.96 | 4.1E-12 | 0.9 | 0 |
| *HLA^b^* | rs9268557 | 6 | 32389305 | T | C | 0.75 | 0.70-0.81 | 4.7E-14 | 0.76 | 0.73-0.79 | 4.5E-47 | 0.89 | 0.87-0.90 | 1.5E-53 | 0.8 | 0 |
| *LY86* | rs12530084 | 6 | 7214676 | T | C | 0.94 | 0.86-1.01 | 9.9E-02 | 0.87 | 0.83-0.91 | 1.3E-09 | 0.95 | 0.94-0.97 | 4.0E-09 | 0.1 | 62 |
| *LYN* | rs75413466 | 8 | 56852496 | A | G | 1.01 | 0.62-1.64 | 9.8E-01 | 1.4 | 1.27-1.56 | 1.4E-10 | 1.16 | 1.10-1.22 | 3.1E-08 | 0.2 | 42 |
| *DEFA1/4^c^* | rs2075836 | 8 | 6808722 | T | G | 1.17 | 1.08-1.28 | 2.3E-04 | 1.21 | 1.14-1.28 | 5.8E-11 | 1.06 | 1.05-1.08 | 1.1E-12 | 0.6 | 0 |
| *CARD9* | rs4077515 | 9 | 139266496 | T | C | 1.1 | 1.02-1.18 | 1.8E-02 | 1.14 | 1.10-1.18 | 2.7E-11 | 1.06 | 1.04-1.07 | 2.4E-11 | 0.4 | 0 |
| *ZMIZ1* | rs1108618 | 10 | 81043743 | A | G | 1.09 | 1.01-1.18 | 2.8E-02 | 1.14 | 1.09-1.18 | 1.9E-10 | 1.05 | 1.04-1.07 | 2.1E-10 | 0.3 | 0 |
| *OVOL1/RELA* | rs10896045 | 11 | 65555524 | A | G | 0.98 | 0.90-1.05 | 5.3E-01 | 1.18 | 1.13-1.24 | 4.8E-13 | 1.05 | 1.03-1.06 | 2.5E-09 | 0 | 94 |
| *ITGAM/ITGAX^c^* | rs10782004 | 16 | 31363417 | A | G | 1.18 | 1.08-1.28 | 1.4E-04 | 1.17 | 1.12-1.21 | 9.4E-14 | 1.06 | 1.05-1.08 | 1.8E-15 | 0.8 | 0 |
| *IRF8* | rs1879210 | 16 | 86017715 | T | C | 1.08 | 0.99-1.18 | 7.3E-02 | 1.14 | 1.09-1.20 | 9.9E-09 | 1.05 | 1.03-1.07 | 1.5E-08 | 0.3 | 15 |
| *TNFSF12/13* | rs3803800 | 17 | 7462969 | A | G | 1.17 | 1.08-1.27 | 2.0E-04 | 1.15 | 1.10-1.20 | 1.2E-10 | 1.06 | 1.05-1.08 | 2.0E-12 | 0.7 | 0 |
| *FCAR^b^* | rs1865097 | 19 | 55397217 | A | G | 1.36 | 1.26-1.46 | 1.1E-15 | 1.12 | 1.08-1.16 | 7.7E-09 | 1.07 | 1.05-1.09 | 2.3E-16 | 0 | 94 |
| *LIF/OSM* | rs4823074 | 22 | 30512478 | A | G | 1.12 | 0.83-0.97 | 4.7E-03 | 0.84 | 0.80-0.88 | 7.8E-15 | 0.94 | 0.93-0.95 | 5.3E-15 | 0.4 | 0 |

a. Novel loci for both phenotypes discovered by cross-phenotype meta-analysis (also highlighted in red).
b. Novel loci discovered in the meta-analysis of IgAV cohorts (also highlighted in blue).
c. Loci that had strong evidence for colocalization between IgAV and IgAN.

**Supplementary Table 12. Colocalization analysis of GWAS loci between IgAV and IgAN.** PP0: probability of no association; PP1: probability of the locus being associated with only IgAV; PP2: probability of the locus being associated with only IgAN; PP3: probability of not sharing the same casual variant at the locus; and PP4: probability of sharing the same causal variant at the locus between the two traits.

| **Locus_name** | **nSNPs** | **CHR** | **Position.A** | **Position.B** | **PP0** | **PP1** | **PP2** | **PP3** | **PP4** |
| --- | --- | --- | --- | --- | --- | --- | --- | --- | --- |
| *INPP5D* | 2688 | 2 | 233715487 | 234514763 | 0.000 | 0.001 | 0.000 | 0.008 | 0.991 |
| *ANKRD55/IL6ST* | 2703 | 5 | 55039706 | 55838612 | 0.001 | 0.000 | 0.018 | 0.007 | 0.973 |
| *ITGAM* | 1356 | 16 | 30968589 | 31762949 | 0.000 | 0.000 | 0.060 | 0.017 | 0.923 |
| *DEFA* | 2575 | 8 | 6408750 | 7007579 | 0.000 | 0.000 | 0.067 | 0.043 | 0.889 |
| *BANK1* | 2929 | 4 | 102388868 | 103188478 | 0.025 | 0.051 | 0.105 | 0.218 | 0.601 |
| *WLS* | 2626 | 1 | 68320888 | 69120590 | 0.026 | 0.005 | 0.449 | 0.088 | 0.432 |
| *TNFSF13* | 1997 | 17 | 7063519 | 7862861 | 0.000 | 0.000 | 0.026 | 0.574 | 0.400 |
| *HORMAD2* | 2214 | 22 | 30119636 | 30918825 | 0.000 | 0.000 | 0.488 | 0.117 | 0.394 |
| *DUSP22* | 1339 | 6 | 188937 | 649516 | 0.000 | 0.000 | 0.547 | 0.064 | 0.389 |
| *CD28* | 2016 | 2 | 204185404 | 204984410 | 0.000 | 0.000 | 0.471 | 0.206 | 0.323 |
| *PADI4* | 2766 | 1 | 17258990 | 18029611 | 0.003 | 0.001 | 0.460 | 0.220 | 0.316 |
| *FCGR2A* | 1977 | 1 | 161071329 | 161869959 | 0.070 | 0.063 | 0.320 | 0.286 | 0.261 |
| *INPP5E* | 2285 | 9 | 138866514 | 139666473 | 0.000 | 0.000 | 0.700 | 0.114 | 0.187 |
| *REL* | 1706 | 2 | 60719970 | 61519408 | 0.000 | 0.000 | 0.256 | 0.567 | 0.176 |
| *FCAR* | 2888 | 19 | 54997421 | 55796482 | 0.000 | 0.003 | 0.000 | 0.981 | 0.016 |

**Supplementary Table 13. Gene set enrichment analysis for 226 IgAV-specific sGenes.** (a) Gene Oncology (GO) enrichment and (b) mouse phenotype enrichment.

| **ID** | **Name** | **pValue** | **FDR B&H** | **FDR B&Y** | **Bonferroni** | **Genes from Input** | **Genes in Annotation** |
| --- | --- | --- | --- | --- | --- | --- | --- |
| **(a) GO enrichment** | | | | | | | |
| GO:0045321 | leukocyte activation | 9.54E-07 | 2.82E-03 | 2.48E-02 | 3.54E-03 | 30 | 1277 |
| GO:0046649 | lymphocyte activation | 2.46E-06 | 2.82E-03 | 2.48E-02 | 9.12E-03 | 26 | 1058 |
| GO:0002682 | regulation of immune system process | 3.92E-06 | 2.82E-03 | 2.48E-02 | 1.45E-02 | 36 | 1821 |
| GO:0050776 | regulation of immune response | 4.08E-06 | 2.82E-03 | 2.48E-02 | 1.51E-02 | 26 | 1088 |
| GO:0002684 | positive regulation of immune system process | 4.53E-06 | 2.82E-03 | 2.48E-02 | 1.68E-02 | 27 | 1164 |
| GO:0002250 | adaptive immune response | 5.28E-06 | 2.82E-03 | 2.48E-02 | 1.96E-02 | 22 | 835 |
| GO:0001775 | cell activation | 5.32E-06 | 2.82E-03 | 2.48E-02 | 1.97E-02 | 31 | 1464 |
| GO:0002764 | immune response-regulating signaling pathway | 7.77E-06 | 3.60E-03 | 3.17E-02 | 2.88E-02 | 18 | 604 |
| **(b) Mouse phenotype enrichment** | | | | | | | |
| MP:0008037 | abnormal T cell morphology | 3.43E-07 | 8.91E-04 | 7.52E-03 | 8.96E-04 | 24 | 835 |
| MP:0012382 | abnormal blood cell physiology | 6.82E-07 | 8.91E-04 | 7.52E-03 | 1.78E-03 | 33 | 1495 |
| MP:0002442 | abnormal leukocyte physiology | 1.12E-06 | 9.75E-04 | 8.24E-03 | 2.94E-03 | 31 | 1379 |
| MP:0001819 | abnormal immune cell physiology | 1.68E-06 | 9.75E-04 | 8.24E-03 | 4.39E-03 | 31 | 1405 |
| MP:0002421 | abnormal cell-mediated immunity | 2.07E-06 | 9.75E-04 | 8.24E-03 | 5.42E-03 | 31 | 1419 |
| MP:0001545 | abnormal hematopoietic system physiology | 2.24E-06 | 9.75E-04 | 8.24E-03 | 5.85E-03 | 33 | 1576 |
| MP:0002420 | abnormal adaptive immunity | 3.04E-06 | 1.14E-03 | 9.59E-03 | 7.95E-03 | 31 | 1445 |
| MP:0002444 | abnormal T cell physiology | 3.92E-06 | 1.19E-03 | 1.00E-02 | 1.02E-02 | 18 | 576 |
| MP:0003945 | abnormal lymphocyte physiology | 0.00000408 | 0.001186 | 0.01002 | 0.01068 | 24 | 960 |
| MP:0006387 | abnormal T cell number | 5.53E-06 | 1.44E-03 | 1.22E-02 | 1.44E-02 | 21 | 777 |
| MP:0002145 | abnormal T cell differentiation | 9.75E-06 | 2.32E-03 | 1.96E-02 | 2.55E-02 | 12 | 284 |
| MP:0008043 | abnormal NK cell number | 1.50E-05 | 3.26E-03 | 2.76E-02 | 3.92E-02 | 9 | 161 |

**Supplementary Table 14. Pathway enrichment analysis of the 773 activated regulators in IgAV cases compared to controls as identified by NaRnEA.** FDR B&H: false discovery rate based on Benjamini–Hochberg procedure.

| **Name** | **Source** | **P Value** | **FDR B&H** | **Genes from Input** | **Genes in Annotation** |
| --- | --- | --- | --- | --- | --- |
| RNA POLYMERASE II TRANSCRIPTION | REACTOME | 5.98E-104 | 1.50E-100 | 251 | 1391 |
| GENE EXPRESSION TRANSCRIPTION | REACTOME | 2.48E-67 | 3.10E-64 | 178 | 1019 |
| GENERIC TRANSCRIPTION PATHWAY | REACTOME | 3.04E-65 | 2.54E-62 | 153 | 766 |
| CHROMATIN MODIFYING ENZYMES | REACTOME | 7.54E-32 | 4.72E-29 | 65 | 272 |
| MECHANISMS ASSOCIATED WITH PLURIPOTENCY | WikiPathways | 9.54E-26 | 4.78E-23 | 61 | 301 |
| SMAD2/3 NUCLEAR PATHWAY | PID Pathways | 6.17E-20 | 2.21E-17 | 29 | 82 |
| HDAC CLASS-I PATHWAY | PID Pathways | 2.18E-19 | 6.82E-17 | 26 | 66 |
| RNA POLYMERASE II PRE-TRANSCRIPTION EVENTS | REACTOME | 9.96E-15 | 2.77E-12 | 24 | 81 |
| TRANSCRIPTIONAL REGULATION BY TP53 | REACTOME | 1.24E-14 | 3.10E-12 | 51 | 363 |
| TRANSCRIPTION OF THE HIV GENOME | REACTOME | 3.32E-14 | 6.94E-12 | 22 | 70 |
| TRANSCRIPTIONAL REGULATION OF WHITE ADIPOCYTE DIFFERENTIATION | REACTOME | 2.11E-12 | 4.06E-10 | 22 | 84 |
| TP53 REGULATES TRANSCRIPTION OF DNA REPAIR GENES | REACTOME | 3.16E-12 | 5.65E-10 | 19 | 62 |
| ID SIGNALING PATHWAY | WikiPathways | 6.66E-12 | 1.11E-09 | 17 | 50 |
| FORMATION OF RNA POL II ELONGATION COMPLEX | REACTOME | 9.31E-12 | 1.46E-09 | 18 | 58 |
| TGF BETA SIGNALING PATHWAY | WikiPathways | 1.95E-11 | 2.70E-09 | 17 | 53 |
| ADIPOGENESIS | WikiPathways | 2.04E-11 | 2.70E-09 | 26 | 131 |
| TGF BETA RECEPTOR SIGNALING | WikiPathways | 3.81E-11 | 4.33E-09 | 17 | 55 |
| SMAD2 SMAD3 SMAD4 HETEROTRIMER REGULATES TRANSCRIPTION | REACTOME | 6.02E-11 | 6.56E-09 | 14 | 36 |
| ADIPOGENESIS GENES | WikiPathways | 6.96E-11 | 7.27E-09 | 26 | 138 |
| HIV TRANSCRIPTION ELONGATION | REACTOME | 7.69E-11 | 7.71E-09 | 15 | 43 |
| TGF BETA RECEPTOR SIGNALING IN SKELETAL DYSPLASIAS | WikiPathways | 1.33E-10 | 1.28E-08 | 17 | 59 |
| NGF-STIMULATED TRANSCRIPTION | REACTOME | 2.13E-10 | 1.98E-08 | 14 | 39 |
| REGULATION OF PTEN GENE TRANSCRIPTION | REACTOME | 2.37E-10 | 2.10E-08 | 17 | 61 |
| HIV ELONGATION ARREST AND RECOVERY | REACTOME | 2.43E-10 | 2.10E-08 | 13 | 33 |
| CIRCADIAN CLOCK | REACTOME | 3.02E-10 | 2.44E-08 | 18 | 70 |
| AP1 PATHWAY | PID Pathways | 3.02E-10 | 2.44E-08 | 18 | 70 |
| PKMTS METHYLATE HISTONE LYSINES | REACTOME | 3.27E-10 | 2.56E-08 | 15 | 47 |
| MYC REPRESS PATHWAY | PID Pathways | 4.15E-10 | 3.15E-08 | 17 | 63 |
| REGULATION OF LIPID METABOLISM BY PPAR-ALPHA | REACTOME | 4.36E-10 | 3.21E-08 | 23 | 118 |
| TRANSCRIPTIONAL ACTIVITY OF SMAD2 SMAD3 SMAD4 HETEROTRIMER | REACTOME | 4.62E-10 | 3.31E-08 | 14 | 41 |
| SUMOYLATION | REACTOME | 9.37E-10 | 6.52E-08 | 29 | 189 |
| INITIATION OF TRANSCRIPTION AND TRANSLATION ELONGATION AT THE HIV 1 LTR | WikiPathways | 2.37E-09 | 1.56E-07 | 12 | 32 |
| EPIGENETIC REGULATION OF GENE EXPRESSION | REACTOME | 3.22E-09 | 2.02E-07 | 28 | 187 |
| ENDODERM DIFFERENTIATION | WikiPathways | 4.33E-09 | 2.65E-07 | 24 | 143 |
| FOXO-MEDIATED TRANSCRIPTION OF CELL CYCLE GENES | REACTOME | 6.03E-09 | 3.60E-07 | 9 | 17 |
| SIGNALING BY NUCLEAR RECEPTORS | REACTOME | 6.19E-09 | 3.61E-07 | 36 | 296 |
| ROLE OF HYPOXIA ANGIOGENESIS AND FGF PATHWAY IN OA CHONDROCYTE HYPERTROPHY | WikiPathways | 8.70E-09 | 4.96E-07 | 14 | 50 |
| ESTROGEN-DEPENDENT GENE EXPRESSION | REACTOME | 1.00E-08 | 5.59E-07 | 24 | 149 |
| ESTROGEN SIGNALING | WikiPathways | 1.17E-08 | 6.38E-07 | 17 | 77 |
| CIRCADIAN RHYTHM GENES | WikiPathways | 1.65E-08 | 8.79E-07 | 28 | 201 |
| MYC ACTIV PATHWAY | PID Pathways | 1.77E-08 | 9.22E-07 | 17 | 79 |
| DELTA NOTCH SIGNALING PATHWAY | WikiPathways | 3.19E-08 | 1.60E-06 | 17 | 82 |

**Supplementary Table 15. The prioritized master regulators in IgAV that were co-modulated by all three GWAS candidate genes (*FCAR*, *INPP5D*, and *IL6R*) in monocytic cell lineage.** pes.values: the raw enrichment score; nes.values: normalized enrichment score; p.values: enrichment P values; fdr.values: false discovery rate; dir.values of UP indicated the increased activity in IgAV patients.

| **Genes** | **pes.values** | **nes.values** | **p.values** | **fdr.values** | **dir.values** |
| --- | --- | --- | --- | --- | --- |
| HIF1A | 0.22 | 12.32 | 6.99E-35 | 5.95E-33 | UP |
| RPL7 | 0.24 | 11.98 | 4.29E-33 | 2.97E-31 | UP |
| ZCCHC24 | 0.23 | 11.91 | 1.07E-32 | 6.75E-31 | UP |
| SMAD3 | 0.24 | 11.63 | 2.83E-31 | 1.53E-29 | UP |
| ZNF219 | 0.22 | 11.39 | 4.95E-30 | 2.54E-28 | UP |
| PHF1 | 0.23 | 11.30 | 1.28E-29 | 6.04E-28 | UP |
| RFX2 | 0.22 | 11.18 | 4.95E-29 | 2.15E-27 | UP |
| CNOT1 | 0.20 | 10.95 | 6.80E-28 | 2.55E-26 | UP |
| ATF7IP2 | 0.24 | 10.93 | 7.95E-28 | 2.88E-26 | UP |
| JARID2 | 0.20 | 10.63 | 2.16E-26 | 6.29E-25 | UP |
| HIC1 | 0.22 | 10.60 | 2.84E-26 | 8.15E-25 | UP |
| TCEAL3 | 0.22 | 10.58 | 3.66E-26 | 1.03E-24 | UP |
| ADNP2 | 0.21 | 10.55 | 5.10E-26 | 1.39E-24 | UP |
| ERF | 0.22 | 10.40 | 2.59E-25 | 6.67E-24 | UP |
| EPC1 | 0.21 | 10.39 | 2.71E-25 | 6.81E-24 | UP |
| ZFP91 | 0.23 | 10.39 | 2.71E-25 | 6.81E-24 | UP |
| GMEB2 | 0.23 | 10.36 | 3.79E-25 | 9.42E-24 | UP |
| ELL2 | 0.21 | 10.29 | 7.89E-25 | 1.90E-23 | UP |
| ZBTB43 | 0.19 | 10.25 | 1.16E-24 | 2.74E-23 | UP |
| GABPB1 | 0.22 | 10.16 | 2.97E-24 | 6.50E-23 | UP |
| IRF2BP2 | 0.20 | 9.89 | 4.51E-23 | 8.50E-22 | UP |
| PPARD | 0.22 | 9.88 | 5.05E-23 | 9.39E-22 | UP |
| AHR | 0.19 | 9.82 | 9.23E-23 | 1.63E-21 | UP |
| NR1H2 | 0.21 | 9.81 | 1.05E-22 | 1.83E-21 | UP |
| BRWD1 | 0.21 | 9.74 | 2.05E-22 | 3.51E-21 | UP |
| MAFF | 0.18 | 9.71 | 2.73E-22 | 4.54E-21 | UP |
| EAF1 | 0.22 | 9.70 | 3.16E-22 | 5.17E-21 | UP |
| MEF2D | 0.21 | 9.49 | 2.42E-21 | 3.67E-20 | UP |
